# Direct Prosthesis Force Control with Tactile Feedback May Connect with the Internal Model

**DOI:** 10.1101/2024.12.05.24318338

**Authors:** Nabeel Hasan Chowdhury, Susan Schramfield, Patrick Pariseau, Dustin James Tyler

**Affiliations:** Department of Biomedical Engineering, Case Western Reserve University, Cleveland, OH, United States; Louis Stokes Cleveland VA Medical Center, United States Department of Veterans Affairs, Cleveland, OH, United States

## Abstract

**Introduction:** Dynamic modulation of grip occurs mainly within the major structures of the brain stem, in parallel with cortical control. This basic, but fundamental level of the brain, is robust to ill-formed feedback and to be useful, it may not require all the perceptual information of feedback we are consciously aware. This makes it viable candidate for using peripheral nerve stimulation (PNS), a form of tactile feedback that conveys intensity and location information of touch well but does not currently reproduce other qualities of natural touch. Previous studies indicate that PNS can integrate with the basic levels of the motor system at a pre-perceptual level and can be processed optimally in multisensory integration, but there is little evidence if PNS is used effectively for motor corrections.

**Methods:** We performed a study with an individual with a mid-radial upper limb difference who has cuff electrodes on his peripheral nerves to give him the sense of touch to perform an object movement over a barrier task. During this task we measured how the participant moved the object with a prosthetic hand in space, how they varied their grip force on the object, and how their muscle signals varied as force changed. We tested this with four different conditions: with and without stimulated tactile sensation combined with the user having control over force on an object or velocity of hand movement.

**Results:** Given direct control of force, the participant’s output force significantly correlated with the puck’s displacement up to the apex of the movement, but did not correlate afterwards. This indicated a trend of increasing force when lifting the puck, but no decrease when lowering it. In comparison, when the participant moved the puck with the intact hand, they had a small but significant increase in force when lifting the puck in half the cases, but always had a significantly decrease in force when lowering the puck. When the participant used a force controller with stimulation, the puck slipped or dropped significantly more times (p < 0.05) compared to the velocity controller with stimulated feedback. This result implied that when the participant intended to loosen their grip, the prosthesis opened instead, which would explain the lack of force reduction in the initial results. The analysis of intent decoded from EMG during use of the force controller shows that the participant intended to lower their grip force with or without stimulation when using a high shatter threshold, but when using a lower threshold, the stimulation gave the participant a better sense of where the shatter threshold was. With a low shatter force, the participant tended to modulate their muscle contractions to a constant level if they were given stimulation (no significant correlation with movement) or they generally increased their intended force towards the shatter force threshold without stimulated feedback. With a moderate shatter force, the participant kept a relatively constant contractile force with or without stimulation. In contrast the EMG analysis with the velocity controller has a mixed trend of increasing and decreasing muscle indicating no global desire to change their grip force in one direction or the other. Finally, analysis of the puck movement showed that the participants moved the puck higher above the barrier with the force controller compared to movements with the velocity controller (p < 0.001), but the addition of stimulation with either controller lowered the participant’s movements significantly closer to the barrier (p < 0.001). Stimulation may cause an instantaneous increase in confidence with a controller or create better positional awareness with either controller.

**Discussion:** While the participant of this study did not show any significant output grip force changes during the object movement tasks, their decoded intent combined with the higher number of loosening events when using the force controller and with stimulation indicates they may have been trying to reduce their grip force during the task. This behavior matches with the force output of the participant’s intact hand. In order convert the participant’s intent into the correct output force, there needs to be changes to the overall design of modern prosthetic devices to allow for smaller grip force changes and changes to force within a static grip. Furthermore, improvements to the stimulation that amplify small changes in force and estimate the any slip forces on the fingertips will provide more useful signals to the participant.

## Introduction

Touch is a direct interface with the world. Other senses like vision, hearing, and smell tell us about the state of the world from a distance, but touch leaves no space between ourselves and the environment. It is for this reason that touch often supersedes the other senses in processing (Caclin et al., 2002; Lunghi et al., 2010; Godlove et al., 2014). However, for individuals with upper limb differences who use prostheses, their direct connection to the world has been severed, forcing them to rely solely on their other senses when using a prosthetic hand. Neural stimulation can create a sensation on the phantom or missing hand over the prosthesis, but it feels unlike receptor generated touch (Graczyk et al., 2018, 2022). Fortunately, the perception of touch is much less important of general object movement. For simple object movement tasks, touch is generally used by the brain stem to change the grip force of the hands automatically as needed without conscious effort (Loutit and Potas, 2020). The brain stem is also likely to be robust to artificial feedback from neural stimulation (Loutit and Potas, 2020) .

Much of the day-to-day usage of touch happens in the brain stem, well before the perception of touch (Libet, 1965, 1993; Abrahams et al., 1988; Imanaka et al., 2002; Henschke et al., 2015; Redinbaugh et al., 2020). At this level, touch is used for automatic corrections to grasp as the weight of objects shift in the hands and if there are any external forces that affect the object (Cole and Abbs, 1988; Johansson and Westling, 1988; Flanagan and Wing, 1993). The many structures of this level of the brain use touch to dynamically modify our actions and combine touch with the other senses to refine our movements without the need for conscious perception. (Ide and Li, 2011; Jang and Kwon, 2015; Risso et al., 2019; Loutit and Potas, 2020; Redinbaugh et al., 2020). Conscious perception of touch is primarily used for novel tasks and experiences, but once those interactions become repetitive, even those interactions become less perceptible (Dehaene and Changeux, 2011). This allows for the limited resource of conscoius awareness to be utilized elseware.

During simple object movements, we respond to the changes in touch much faster than should be possible with conscious decision making. For example, multiple studies show that we modulate grip force to oppose the inertia and changing load forces on an object within 60-90 ms (Cole and Abbs, 1988; Johansson and Westling, 1988; Flanagan and Wing, 1993). Reactions of this speed occur in the simplest areas of brain well before touch is consciously processed (Scott, 2016). For comparison, the fastest humans can consciously respond to a tactile stimulus is on the order of 200 ms and fastest to respond to a visual stimulus is 300 ms or more (Godlove et al., 2014; Woods et al., 2015). With grip modulation occurring at a third to even a fifth of this time, dynamic modulation of grip must occur without conscious intervention.

There are many parallel structures of the brain stem where tactile feedback augments grip control (Loutit and Potas, 2020), but one of the best understood is the cerebellum. Though there is debate on the internal connections of touch within the cerebellum, the general consensus is that the cerebellum receives descending connections from the somatosensory cortex about the expectations of what a movement will feel like and ascending connections from the periphery as the truth of what the motion did feel like (Snider and Stowell, 1944; Thach, 1998; Manni and Petrosini, 2004). The current accepted theory for how these streams of information connect is in a massively parallel set of cerebellar microcircuits. This network of circuits has two purposes. First, the cortex uses the internal model of the body to generate the expectation of how the movement will feel. This lets the cerebellum filter out any self-generated tactile feedback so that only the new or unexpected tactile information is used in further processing (Latash, 2021). Second, the unexpected tactile feedback is used as a reference of what actually happened as a result of a motor action. The cerebellum uses this error signal to make minor corrections to the motor signals descending to the spine in order to bring new sensory signals closer to the body’s expectation as well as update the body’s internal model. This is what is called the adaptive filter model of the cerebellar microcircuit first proposed by Fujita and used today to quickly train robots to make fewer mistakes (Porrill et al., 2013; Wilson et al., 2015; Fujita, 2021). The cerebellum is essentially a self-contained autonomous brain within the larger brain used for perception and decision making. The cerebellum and the brain stem nuclei reduce the amount of conscious processing done by the higher levels of the central nervous system by automatically processing sensory information and adjusting motor actions (Loutit et al., 2020).

For those who have lost their hand, many still have a general perception of their hand called the phantom limb. Stimulation of the somatosensory cortex can sensation on the hand (Bensmaia, 2015; Caldwell et al., 2019), which indicates that the body’s mapping of the missing hand still exists even if there are no receptor generated tactile signals coming from the hand. Typically, those with limb differences must now use their other senses to substitute for touch (Chadwell et al., 2016), but these substitution methods require the person to consciously refer the sensation to its meaning. Sensory substitution in this manner would not take advantage of the existing automatic processing of touch in the cerebellum. For this, a signal needs to ascend through the existing neural pathways for touch.

Stimulation of the peripheral nerves that once connected to the hand evokes tactile sensation on the phantom limb (Tan et al., 2014). While the perceptual information of this stimulation is generally limited to location and intensity, the perceptual qualities of touch are less important for basic, automatic touch processing and usage (Loutit and Potas, 2020). For example, peripheral nerve stimulation (PNS) integrates optimally with the other senses (Risso et al., 2019). This integration mostly occurs in the brain stem before perception. Our previous work shows that PNS is processed as fast as naturally generated touch and contributes to motor control without the need for perception of the stimulation (Chowdhury and Tyler, 2024). PNS can also elicit the spinal H-reflex in intact participants (Pierrot-Deseilligny et al., 1981; Fung and Barbeau, 1994) and can modulate some spinal reflexes (Dalrymple et al., 2024).

These studies suggest artificial touch produced by PNS can be processed by the brain stem, but it is not known if PNS is actively used in dynamic motor control changes. The basic processing in the earlier studies requires little information other than the signal is a touch sensation. Grip control may require more information aside from location and intensity. However, studies show that stimulated touch both in the periphery and directly to the cortex improve functional performance with prosthetic and robotic devices (Tan et al., 2014; Graczyk et al., 2018; Flesher et al., 2021). These functional improvements can occur instantaneously with the addition of touch and disappear with its removal (Flesher et al., 2021). They also can increase over time given prolonged usage at home and then reduce once the artificial touch is removed (Graczyk et al., 2018). In the former case, stimulation seemed to give the participant a better understanding of their robotic hand which shows perceived touch can create immediate functional benefits. This study was performed with a spinal cord injury survivor so, it is not possible to say if the same benefits will occur with a prosthesis user. In the latter study, touch ascending through the tactile path seemed to be learned over time which increased scores on standardized functional measures. The tests in this study however required a focus on what the participant was feeling during the task and the tasks were more complex than a simple object movement. There were no recorded metrics on the integration between touch and the motor system.

Simple object manipulation uses tactile feedback well below the level of perception to make minor corrections to grip. This study examined the integration of PNS tactile feedback in the pre-perceptual neuromuscular control pathways. An individual with a transradial limb difference and a CFINE electrode on his median nerve participated in the study. The object that was moved in the test has a sensor to measure grip force and the participant used either their original velocity-based prosthesis grip controller or a custom force-based grip controller.

Given direct control of force on an object and stimulation, we hypothesized that the participant’s output force on an object will correlate with the motion of the object in space. The results of this study show that given direct control of force, the participant increases force when lifting the puck and intended to decrease force when lowering it.

## Methods

### Research Participant

The participant of this study is a left, transradial amputee who lost their arm due to a traumatic injury. He was implanted with recording and stimulating electrodes as part of multiple studies and participated in this study over the course of a year. For this study, he performed 15 sessions of between 10 to 30 trials over the course of one to two hours depending on his availability and came in about once a month. This resulted in a total of 479 total trials recorded.

### Implanted Electrodes

The research participant has 2 Composite Flat Interface Nerve Electrodes (CFINEs) (Dweiri et al., 2016; Freeberg et al., 2017) implanted around their median and ulnar nerves and 8 intramuscular electrodes in the Pronator Teres, FCR, FDS, FCU, Supinator, ECRB, EDC, and ECU. These eight muscles form a set of 4 flexor and 4 extensor muscles.

### Sensorized Puck Movement Task

A participant moved a puck over a barrier from one set location to another and then back. The locations were 11 inches (27.94 cm) from the barrier on each side and the barrier was 20 cm high (Figure 1). The barrier height and locations were based on the box and blocks test (Mathiowetz et al., 1985) where a participant moves as many small, wooden blocks over a barrier as fast as they can without any regard to the starting or end locations. In this test, we asked the participant to be much more controlled in their movements and the speed did not matter. They were instructed to move an instrumented puck from one set position to another in whatever way that was natural to them.

**Figure 1:**
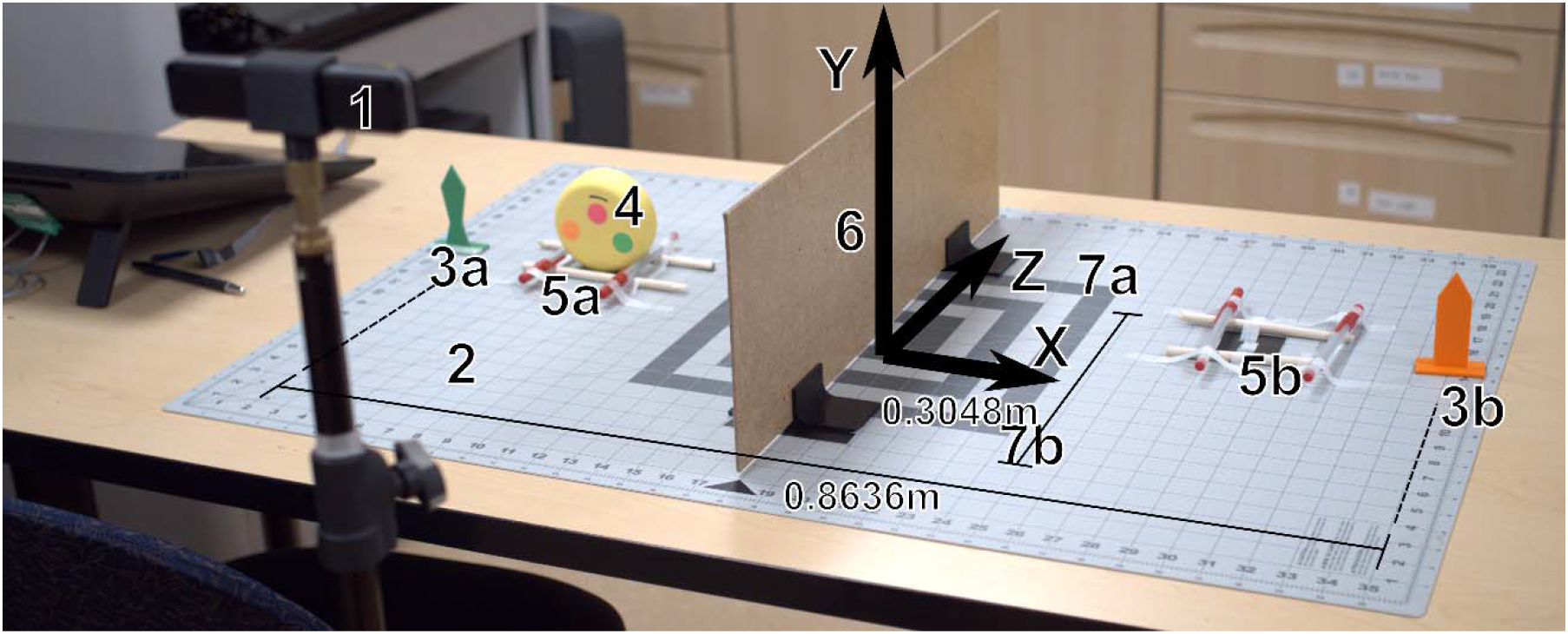
The puck movement tack with parts labeled. Experimental setup with the Zed Mini (1) included. All parts of the setup were in reference to a large crafting mat (2) with pre-marked locations for all testing parts. The participant moved the puck (4) over the barrier (6) between each cradle (5a, 5b). The green, orange, and pink stickers were used to track the puck in space and the orange and green static markers on the mat (3a, 3b) and the corners of the large square on the mat (7a, 7b) gave the DeepLabCut known reference points later used to convert from arbitrary units to real world units. The locations of the static markers were used as reference points to create a new coordinate frame for the movement of the puck in real world units. The distance between the orange and green static markers was 0.8636m. The line these two markers made was the x axis. The distance between the front and back reference points was 0.3048m and the line between them was the z axis. The center point of these two axes was the origin and the direction orthogonal to these upwards was the y axis. Note that the y axis here should be the negative y based on the directions of the x and z axes. The y direction was flipped in the code to make all movements in the positive direction.

The experiment occurred over a grid with positions for all components of the experimental set-up. The experimental set-up (**Figure 1)** consisted of a mat and barrier for the participant to move the puck over. The mat also had designated locations for the puck and other static tracking points. The motion was recorded by a stereoscopic camera called the ZED Mini ^TM^ (StereoLabs, San Franciso, USA).

During the test, the participant was instructed to wait for a verbal signal to start. After the go signal, he grabbed the puck from the first position on his right (**5a**), moved the puck to the designated cradle to on his left (**5b**), let go, and paused shortly. Then the participant grabbed the puck again without prompting and moved the puck back to the original cradle over the barrier. This test was repeated 5 times for each condition of the test for ten movements each. The conditions of the test were with a force-based prosthesis controller or the subject’s original velocity-based controller pair with stimulation being supplied or no stimulation. The control case was with the participant’s contralateral intact hand.

The stimuli during the tests were felt on the participant’s thumb and index finger and the intensity corresponded to grip force measured by the puck. The intensity range of the stimuli was from the threshold of perception to a level the participant reported as feeling like he was squeezing a wooden block as hard as he could. For reference they were given a small wooden block to squeeze with their intact hand. Stimuli in these trials used a standard charge balanced, biphasic wave with a 2:1 ratio of cathodic pulse amplitude to anodic pulse amplitude. The three parameters we could control were the pulse amplitude, pulse width, and pulse frequency. We chose to modulate pulse width to control intensity and leave the other parameters at constant values determined at the start of each experimental session. The top of the range of pulse widths was 250 microseconds and the set pulse amplitude that felt like the participant’s max squeezing force. The threshold was that same pulse amplitude with the minimum pulse width just before the participant stopped feeling the stimuli. The force the participant applied to the puck scaled linearly with the increase in pulse width of the stimuli.

After the first few trial sets, we noticed that the participant was gripping the puck with the full strength of the prosthetic hand when moving the puck to guarantee that they would not drop the puck. We countered this behavior by adding a software defined “shatter” force threshold. Above this force, a sound played to tell the participant that they failed the trial and to start over. The first few attempts at setting a reasonable shatter force varied. The set shatter force for the first three trial dates was so high, 106.36 N, that the prosthesis in the study was no capable of reaching it. Subsequently we measured the prosthesis’s maximum force to be 50.54 N. On the next trial date, we measured the minimum force the intact hand used to lift the puck, 7.41 N, and set the shatter force just under twice this value, 15.02 N. Unfortunately, this shatter force was so low most trials involved a shatter. All tests after this minimum shatter force used three times the intact hand’s minimum force, 22.64 N, which the participant found difficult to stay under by the task was still achievable.

### Force Controller

Our muscles output contractile forces that apply torque to our joints, yet traditional prosthesis controllers convert muscles signals to velocities. We developed a force-based prosthesis controller that converts the participant’s muscles signals to force, because we hypothesized it would provide a more direct connection from the participant’s grip force intent to the prosthesis’s grip force output. Our hypothesis was that the participant would need direct control over force to correctly connect the artificial tactile feedback to the motor system. The force controller in this study was a 1 degree of freedom controller for the grip position of a TASKA hand with sensorized fingertips (TASKA Prosthetics, Christchurch, New Zealand). The force was estimated from the participant’s electromyographic signals (EMG), or the electrical signals their muscles generate when contracting. The EMG was converted to force in a series of steps.

Each EMG signal is rectified and smoothed over a sliding window of 200 ms that slide in 50 ms increments **(Figure 2**).

**Figure 2:**
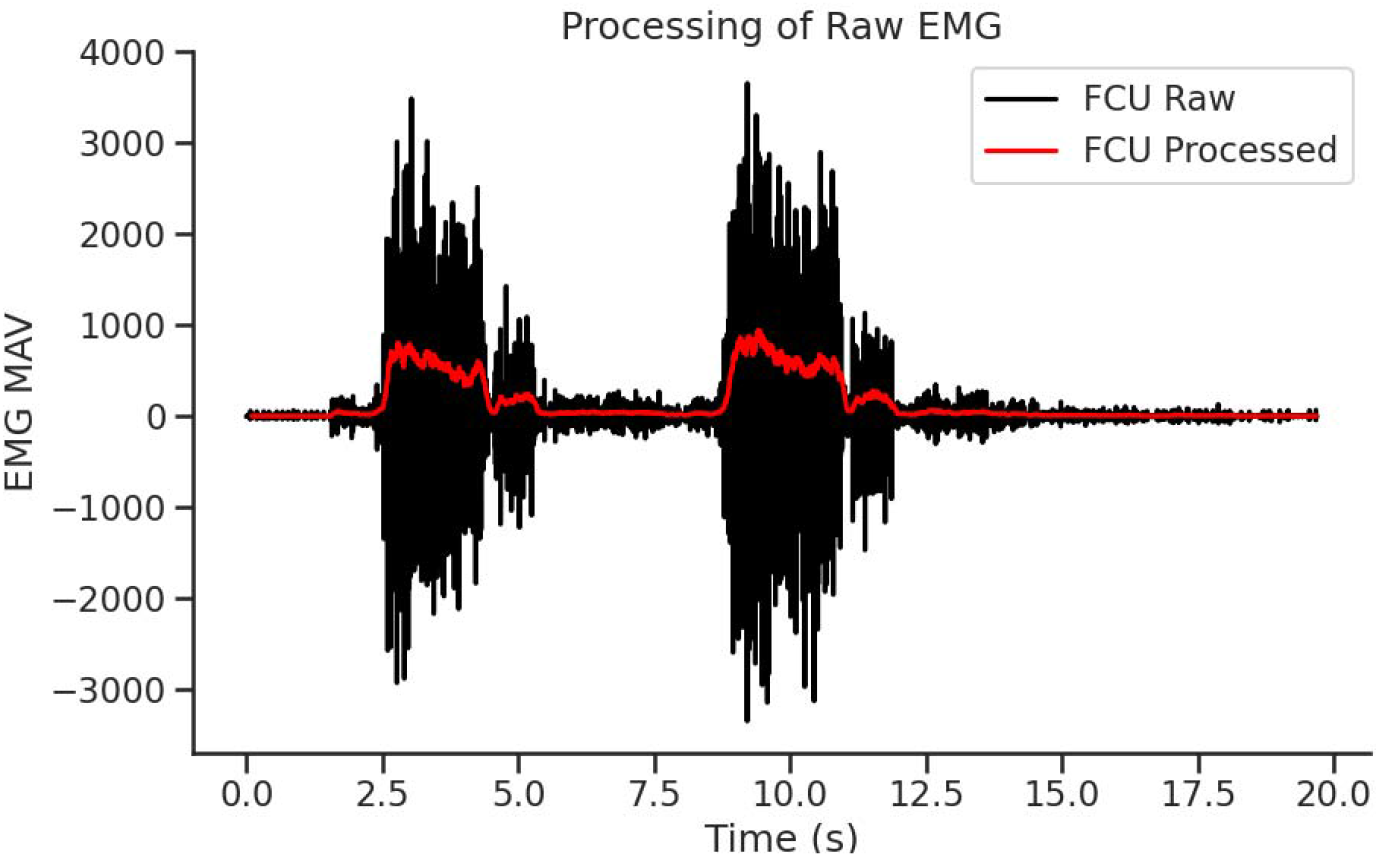
Example processing of raw EMG into each muscles force intent. The raw EMG was window averaged over 200 ms and the window slid every 50 ms. Each channel was then normalized to the muscle’s maximum contraction.

Each signal was normalized from 0 to 1 and the flexor and extensor muscles were combined to form a closing and opening force value. The closing value minus the opening value formed a set point for an average force on the fingertips. The average of the sensor values of the thumb, index finger, and middle finger formed the “real force” on the object and the subtraction of the set point and the “real force” formed the error signal to the controller.

This error signal was used by a PID controller tuned with the Ziegler–Nichols method (Ziegler and Nichols, 1993) to change the aperture size of the prosthesis until a certain force was reached. The maximum change of aperture size was limited to a maximum of 12 out of 255 units every 50ms to slow the overall speed of the hand before it met an object and to limit the error signal in the PID controller from over accumulating. This tuning method eliminated oscillations while also allowing the hand to move fast enough for the participant’s preferences. The validation of the controller tuning is in **Supplementary Figure 1, Supplementary Figure 2, and Supplementary Figure 3**.

### Instrumented Puck

The object moved in this task (**Figure 3**) was a FitMi Motion Interface from Flint Rehab (Flint Rehab, California). The puck has multiple sensors to record force, linear and angular acceleration, and the magnetic field of the earth. For this study, we used the force sensor alone and captured the position of the puck externally to limit the amount of data that needed to be transferred over Bluetooth. The puck force values vary from 0 to 1000 with a baseline at 400. Values below 400 correspond to pulling the faces of the puck apart. The puck’s range of measurable values to are from -101.7 N to 152.03 N (**Supplementary Figure 4**).

**Figure 3:**
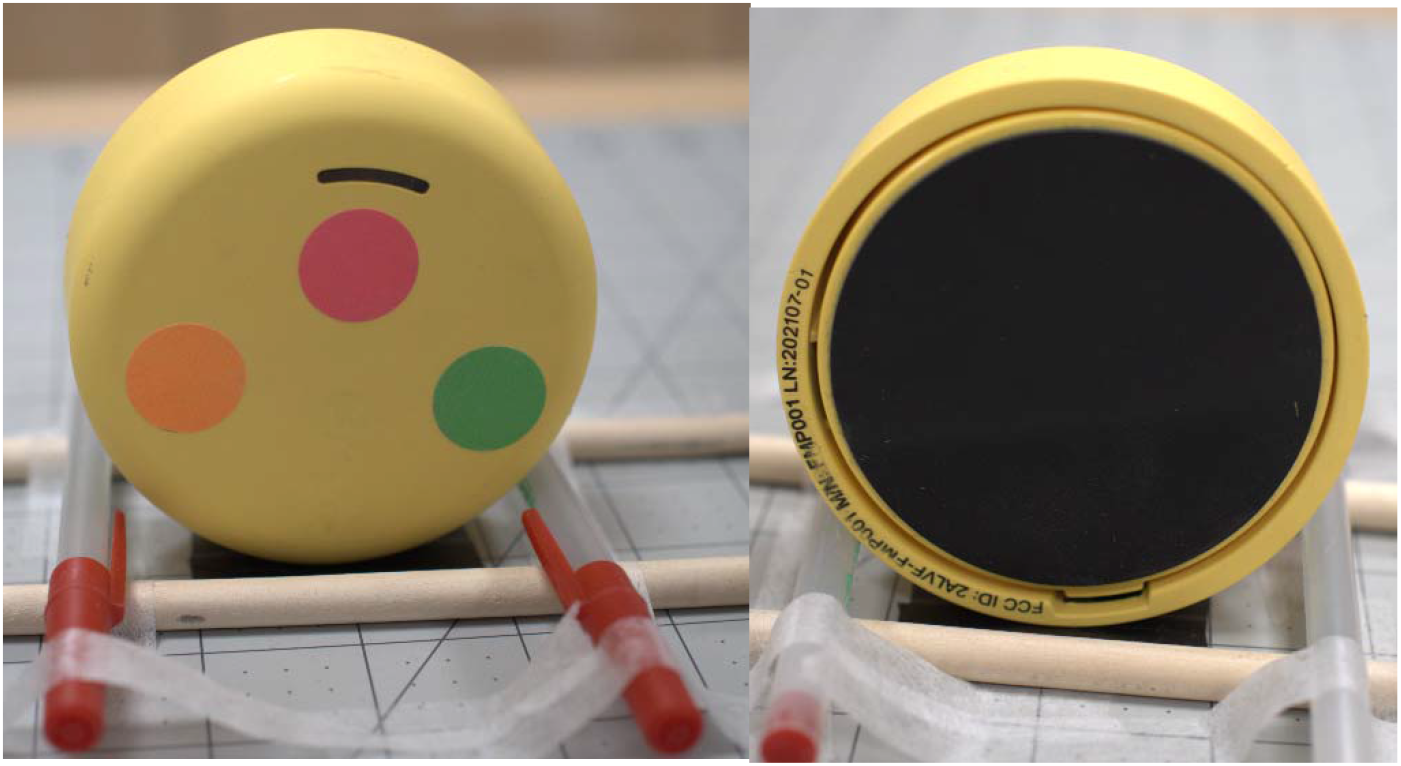
The Flit Rehab FitMi Motion Inteface. The sensorized puck used in the task sitting in the start cradle for the test. The front of the puck (left image) has three colored markers used in motion capture. The back of the puck (right image) has a rubberized surface to aid in grip and shows the space between the front and back faces of the puck. Squeezing these two faces activates the force sensor embedded in the puck and also turns on the light on the front face of the puck.

### Motion Tracking

The puck was tracked using DeepLabCut, a tracker-less motion capture system (Mathis et al., 2018), smoothed by a Kalman Filter (Kalman, 1960) and a kernel convolution on the radial distance from the camera, and aligned by fitting each motion trace to a parabola then aligning the peaks of those parabolas.

### DeepLabCut Motion Capture

We chose DeepLabCut over a commercial motion tracking setup because it easily fit into our existing testing setup without the need to move to a larger room with commercial motion capture. DeepLabCut (DLC) uses a trained neural network to identify and track targeted features in video data of rapid moving participants in varying environments without the need for special markers (Mathis et al., 2018). This network’s robustness minimized the effects of changes in the participant’s appearance between experimental sessions. Despite DeepLabCut not needing trackers, we added green, pink, and orange stickers to the puck to aid in tracking. This allowed us to create an accurate network with few training videos. Training data for the network consisted of sample videos with various people in changing outfits performing the test. This helped train the network to adapt for any outfit worn by the participant and accommodate his prosthetic and intact hands.

Videos of the participant were loaded into the trained network for analysis. Each day data was collected, we recorded an additional calibration video of a checkered grid moved around the experimental workspace. The calibration video helps DLC calculate the distance of the puck from the camera, helps to eliminate error from slight differences the placement of the camera day to day, and minimizes lens distortion.

Pilot data videos recorded by the ZED Mini were HD and at 30 Hz. The rest of the videos were VGA resolution and 100 Hz. DLC outputs tracking data as coordinates points on X, Y, and Z axes in units determined by the pixel count of the source video. We converted these arbitrary units into real world units using the known distance between the static markers common in the experiment trials. The orange and green static markers (**3a** and **3b** in **Figure 1**) were 86.36cm (34”) apart, and the distance from each marker tip to the tabletop is always 7.62cm (3”). The front and back left static markers (**7a** and **7b** in **Figure 1**) were used as Z axis calibration points. The distance between these markers is always 30.48cm (12”). All points were marked in inches because the grid used in the experiment was made in inches (**2** in **Figure 1**).

Because the orange and green static markers were equidistant from the barrier, the center point between them was set as the origin of the x axis. The DLC coordinate system was then rotated around the Z axis until both markers were on the X axis, giving them both a Y coordinate of 0. The true distance between the orange and green marker tips (0.836m) was divided by the distance on the arbitrary DLC coordinate system, and then each data point was scaled by the result and offset by the new origin coordinates. To reduce the amount of “jitter” in the final data, all calculations were performed for each frame of each video.

Tracking errors in DLC occur when the network misidentifies a point in the frame as a tracking marker. This occurs most often when the actual tracking marker is not present in the frame, such as if it is covered by the participant’s hand or if the puck has been dropped. We filtered out moments with obscured tracking markers and drops under two conditions. First, any puck location outside the real-world limits of the testing zone were removed. The distance between the barrier and the edge of the testing mat measures 0.4572 m. Any tracked coordinates more than 0.75 m to the left or right of the barrier were deemed tracking errors, as the participant never took the block more than 0.30 m beyond the edge of the testing zone. Second, any data describing the puck as more than 1 m above the origin was discounted as this distance exceeded the participant’s range of motion. The z range was expanded all the way to 10m because although the testing zone is only 0.6096m deep, measurements on the z axis were highly variable. This noise was dealt with by smoothing of the depth measurement on the processed data described below. This smoothing is of little effect on the finished data as all motions were simplified to a plane orthogonal to the depth axis which flattens all depth measurements.

### Kalman Filter, Spherical Data Smoothing, and Data Alignment

The output of DLC were fed into a Kalman Filter with each sticker on the puck refining the filter’s estimate at each time step. Of the original 479 trials, the first 80 were used as pilot data in tuning the final system and improving upon the experimental design. In the remaining 399 trials, 50 were with the intact hand for comparison to the prosthesis performance and 349 were with the prosthesis. Tracking was largely lost on 95 of the prosthesis trials, and on 11 of the intact trials due to the participant covering the markers on the puck with their hand during the entirety of a trial. A summary of this is in **Table 1**.

**Table 1:** The number of movement trials with and without motion tracking. Of the original 479 trials, the first 80 were used as pilot data, 50 were recordings of the intact hand, and 399 were recordings of the prosthesis. Tracking was lost on 11 of the intact trials and 95 of the prosthesis trials.

|  |  |
| --- | --- |
| Pilot Data Trials | 80 |
| Prosthesis Trials With Tracking | 254 |
| Prosthesis Trials Without Tracking | 95 |
| Intact Hand Trials With Tracking | 39 |
| Intact Hand Trials Without Tracking | 11 |
| Total Trials | 479 |

Once filtered, we saw large jitter in the depth measurements of the puck (**Figure 4**). This should have been accounted for by the calibration step outlined above, but the small distance between the lenses, 6.3 cm, on the ZED Mini resulted in calibration errors.

**Figure 4:**
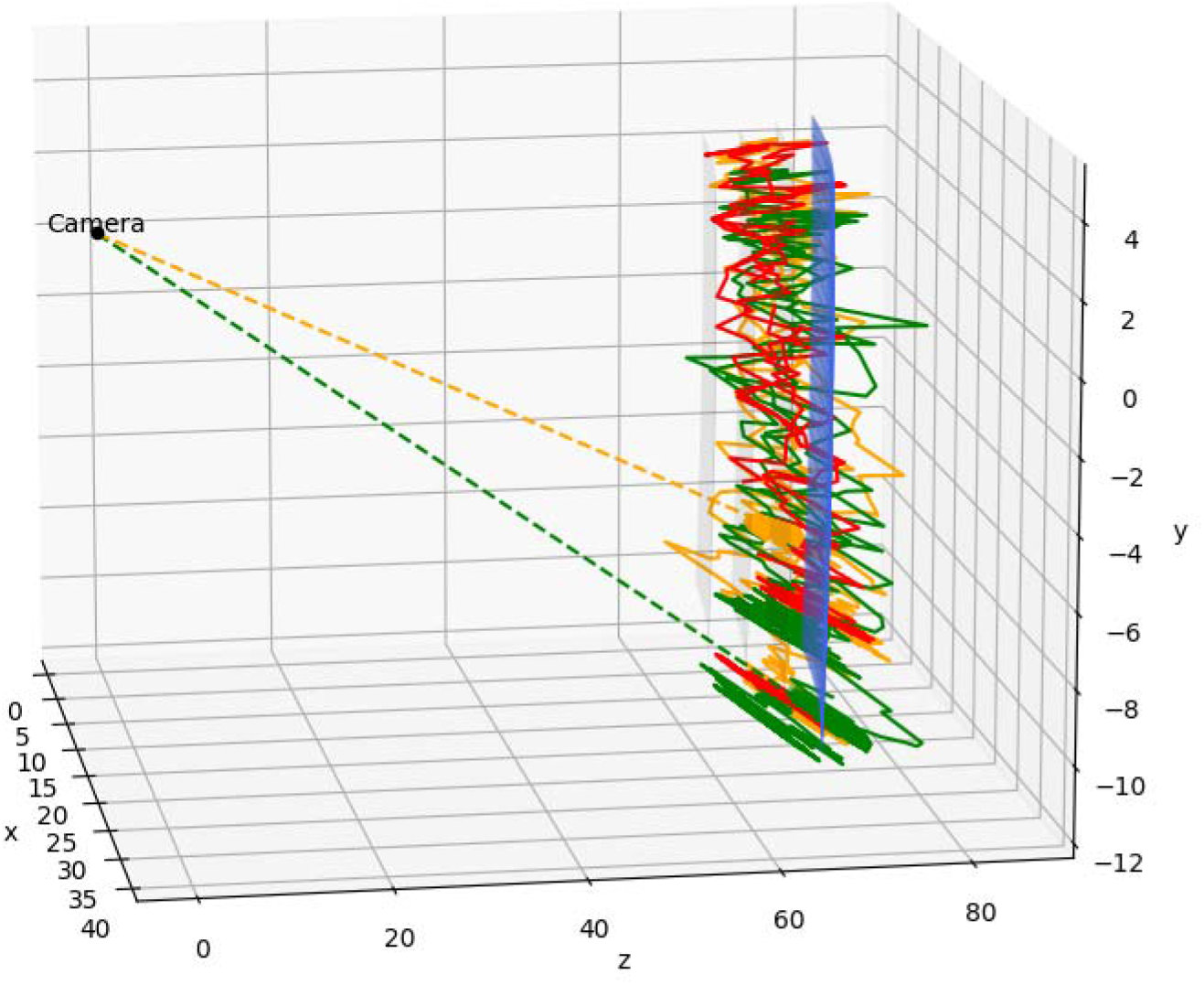
The positions of the green, orange, and red markers jittered radially away from the camera position.

Upon analysis of the noise in the raw DLC recordings, we found that the noise was moving radially away from the camera’s location at the origin. The noise was error in estimates of distance from the camera. Because this noise could be simplified as noise radially away from the origin, we converted the position data to spherical coordinates and smoothed the radial value for each tracking marker with a kernel convolution of 15 points. Then the average position of the red, green, and orange markers was used at the final movement trace. The result of this smoothing and averaging is shown in **Figure 5**. Note the two black traces that represent a movement right to left and left to right. These two movements are one trial.

**Figure 5:**
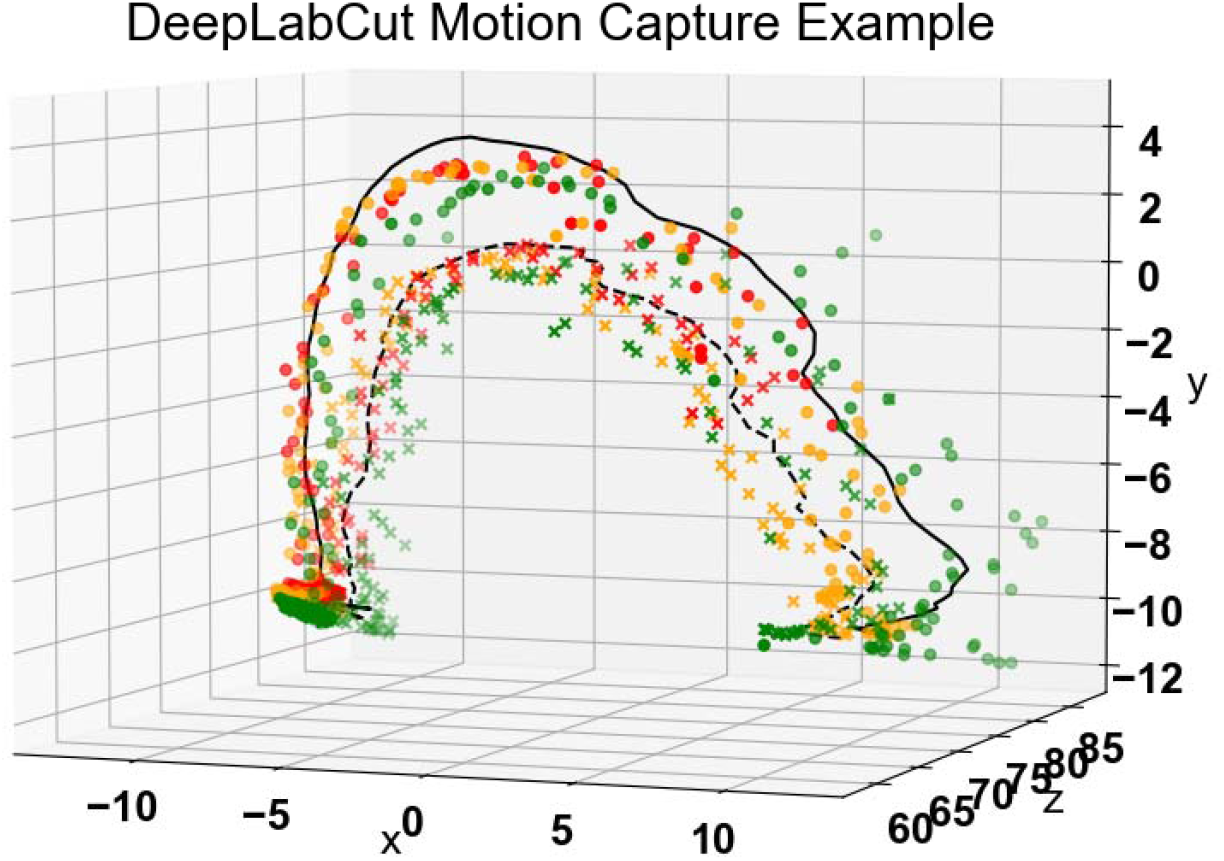
Spherically smoothed movement trace. Data is in pixel units output by DeepLabCut here but the full dataset used real world units in cm. The red, green, and orange markers are each found position of those tracking markers in each video frame. Circles are right to left movement and cross markers are left to right movement positions. The black trace is the average position of the markers in the movement right to left (solid) and then left to right (dashed).

After smoothing, each individual motion trace had different locations within the coordinate frame. This was due to the shifts in the camera’s location day to day. We aligned the trials by fitting each trace along the X and Z axes to have a uniform depth and movement peak location. The height of the traces was unaffected by alignment. Of the 254 traces with a prosthesis, only 30 did not have a fit with an R^2^ value above 0.6 (**Figure 6**) with those traces comprised of major tracking errors where the puck was largely missing in the tracking (**Figure 6**). The rest of the data below is from the 88% of traces that had tracking and could be aligned.

**Figure 6:**
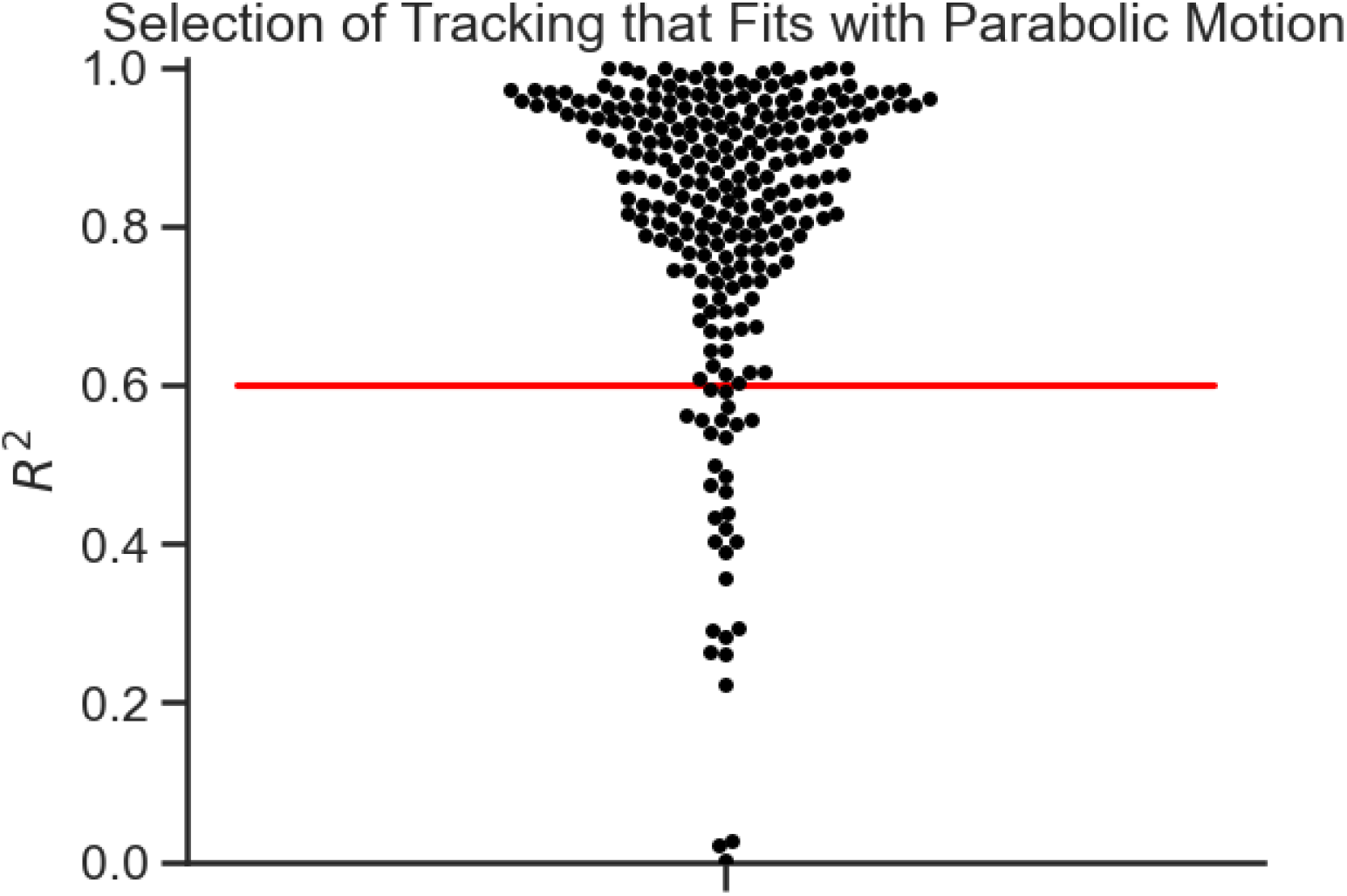
R squared values of all parabolic fits to the movement traces. Above 0.6 was considered a good fit.

The aligned traces are in **Figure 7**. This alignment removed the shifts in each trace position due to the camera shifts day to day and put each trace on a similar global coordinate frame.

**Figure 7:**
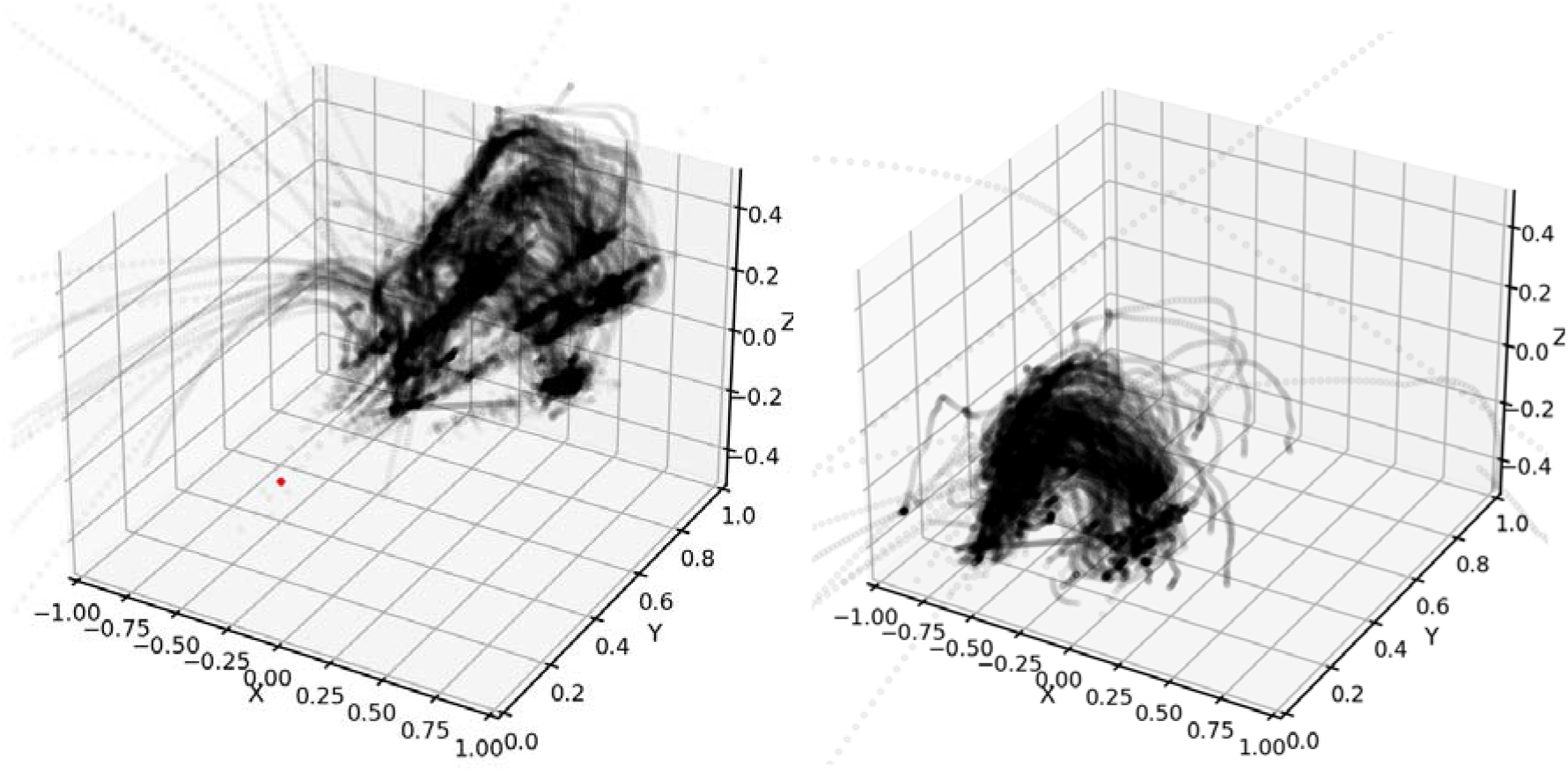
Alignment of the movement traces uses the parabolic fits of each trace to have it peak over the origin. The left figure shows all of the traces before alignment and the right figure shows all traces after alignment.

### Synchronizing the Recordings

There were several different recordings for this experiment which all needed a common recording time. The puck recordings were sent over Bluetooth to the stimulator computer. The received force values were used to stimulate and were recorded with the EMG values on the clock for the stimulator. The DeepLabCut recordings were aligned manually with a sync time indicated by a light on the puck that turned on when 4.9N (or 500 grams) was applied to it. Any attempt to grab the puck would send a force signal to the stimulator computer and the sync time with DLC was the first large and rapid increase in the puck’s force value (**Figure 8)**.

**Figure 8:**
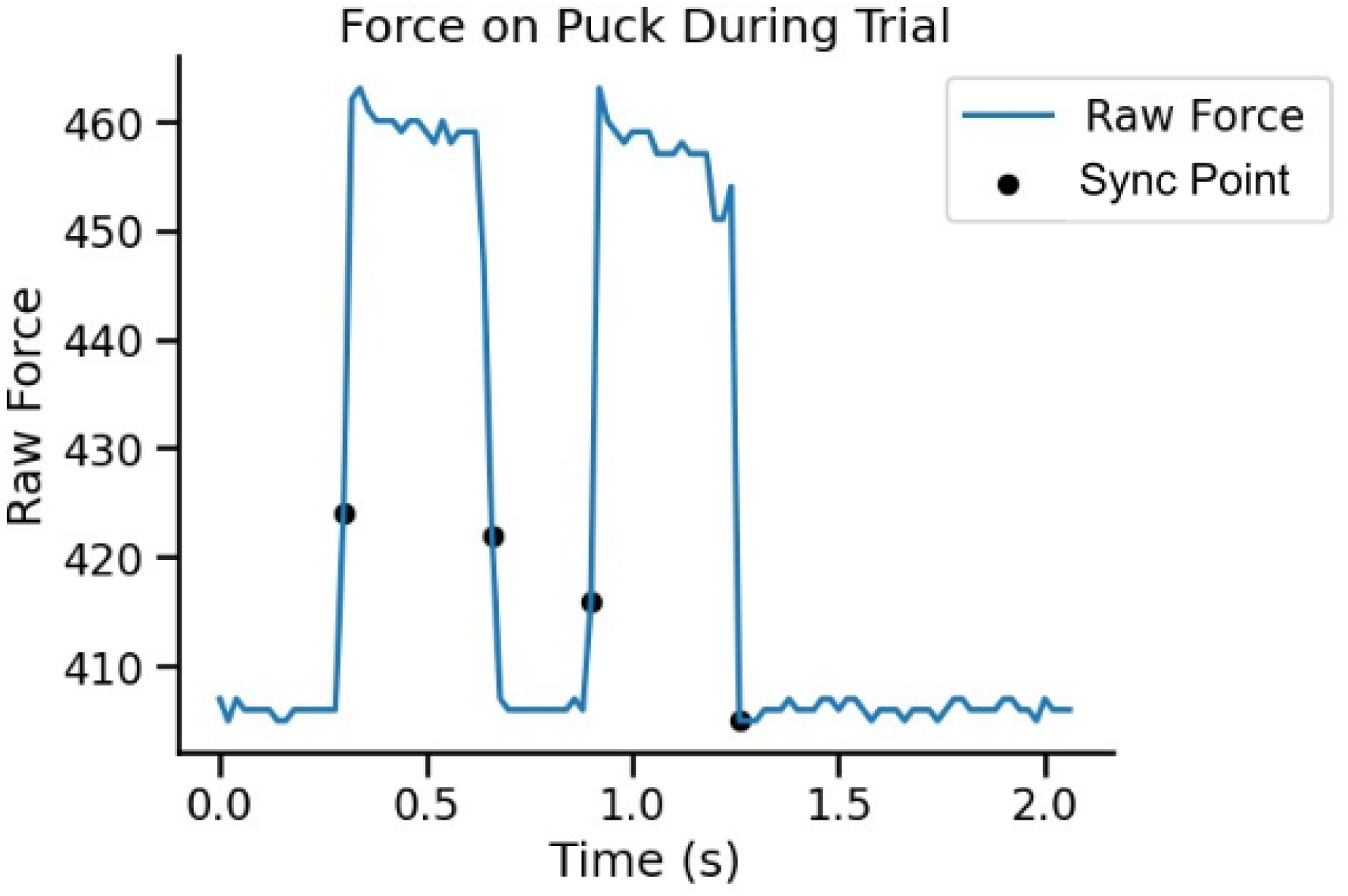
Puck forces. An example force trace with possible sync points. The sync point of the force recordings to the DeepLabCut recording was the first large increase in the force trace recorded by the puck.

## Results

### Force Modulation During Movement

During the puck movement task, the main forces on the puck are gravity and the inertia. The participant uses their grip force and the friction of their prosthesis’s fingertips to oppose these forces. Assuming the force of gravity is constant during the movement of the puck and always points downwards, gravity and the inertial force will oppose the puck’s movements together on the upward movement and inertia will oppose gravity on the downward movement. As a result, we expected that given direct control over the force on the puck, the participant’s grip force would increase towards the apex of movement as they fight against both inertia and gravity and then decrease on the path towards the table with the assistance of gravity. The correlation of puck force to vertical displacement in **Figure 9** shows that on the upward movement with the puck, the participant’s grip force increases, but only when they use the force controller. The positive correlations are also stronger when the force controller has no stimulation added to it. The velocity controller does not have a significant positive correlation with the puck’s movement. In contrast however, on the correlations with downward movement in **Figure 10**, there are fewer significant correlations with the force controller suggesting that the puck was held at a constant force on the downward motion. Missing correlation values were from sparse motion capture data for specific section of the tracking.

**Figure 9:**
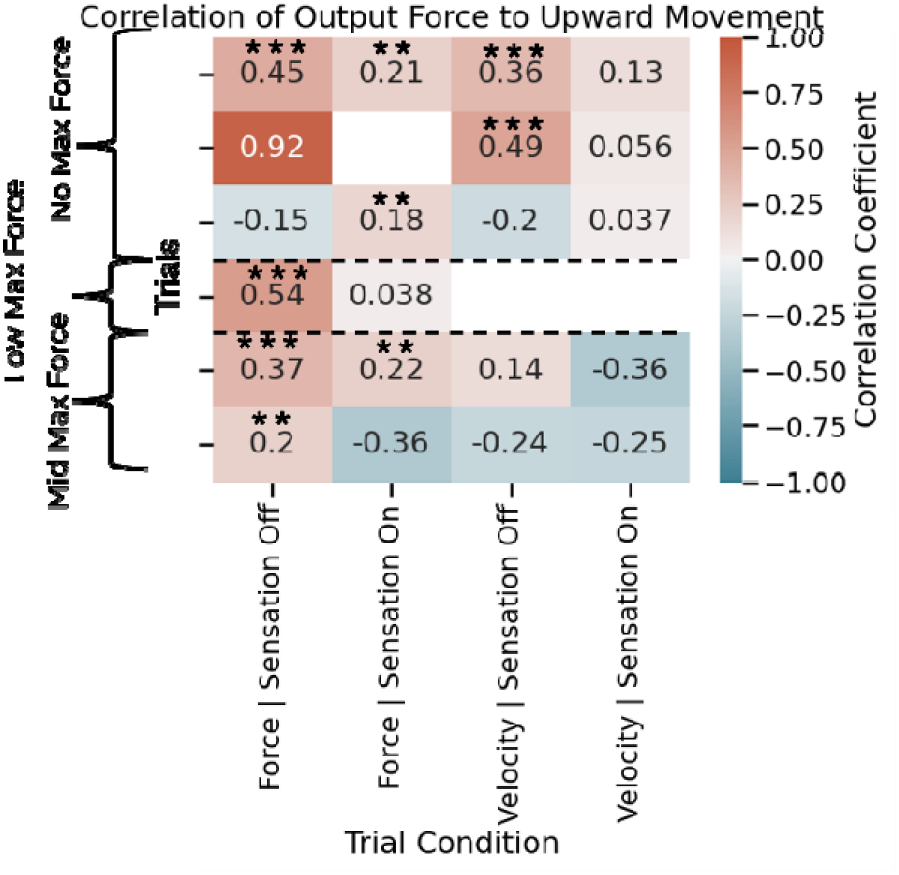
The correlation coefficients for output force to puck upward displacement. Each row is a different trial, and the trials occurred in order from the top row to the bottom row. The participant had a significantly positive increase in output force while lifting the puck with the force controller with or without stimulation, but not with the velocity controller.

**Figure 10:**
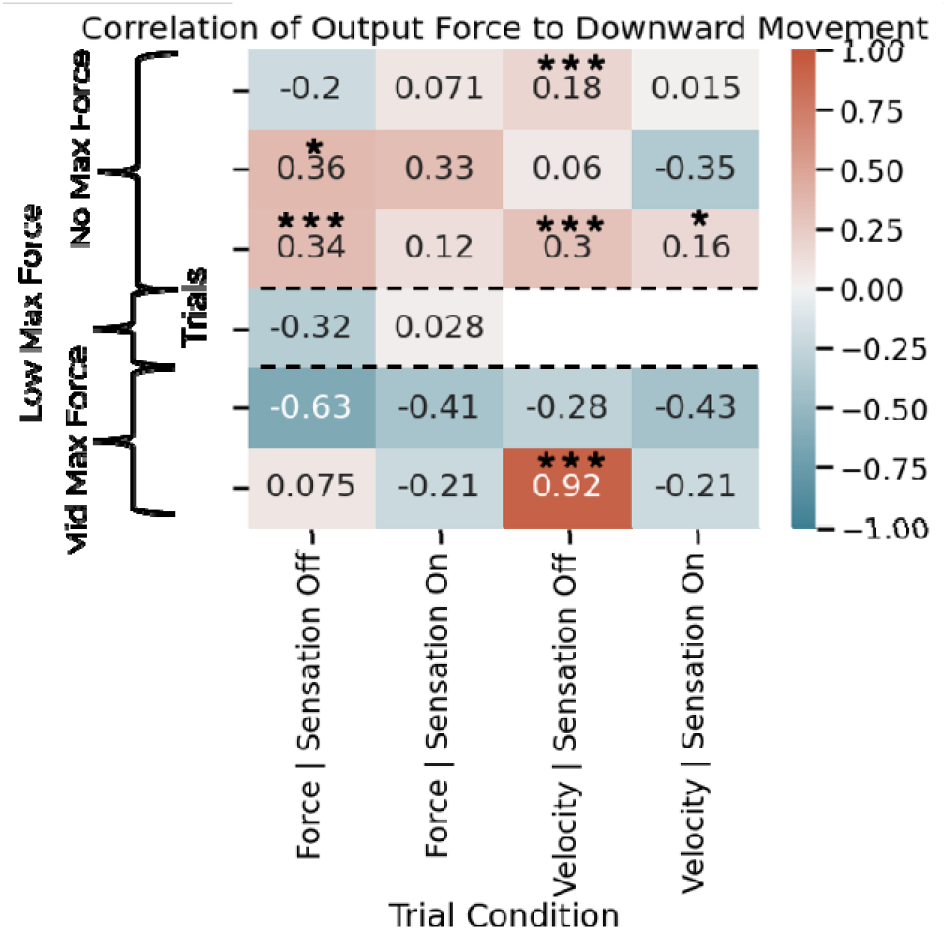
The correlation of output force on the puck with displacement on the downward movement. Each row is a different trial, and the trials occurred in order from the top row to the bottom row. The significant correlations are much sparser compared to the upward movement indicating the participant had a relatively constant force.

The main conclusion to draw from these figures is that the participant’s use of a force controller results in them increasing their force through their movement to the puck’s apex, but they do not seem to reduce their output force on the puck. One reason for this is possibly due to the design of the prosthesis. Most modern prosthetic hands do not have the ability to reduce their grip force on an object while keeping a static position. This would require a prosthesis to be backdrivable or able to modulate its stiffness within a grip position. The result of this appears to be a higher number of slip and drop events seen in the results later as the participant attempts to reduce force.

When the participant performs the task with their intact hand, we see clear trends in correlations aligned with our expectation. For the intact hand trials, the subject unfortunately covered the tracking markers with their hand. This prevented a recording of an accurate displacement, but we can approximate where the subject was moving the puck in space by dividing the puck’s force traces by reference points in the movement. The forces were divided using the manually recorded times of when the puck was grabbed and set down for each trial. The participant had no errors when using their intact hand. This resulted in a consistent movement that was easily split using this technique. Each trace was divided in half with the first half being the ascending portion of the trace and the second half being the descending portion of the trace. The trial time was substituted for the puck displacement and started at zero for each trace section. **Figure 11** shows the participant increased their grip force on the upward movement and decreased their force on the downward movement as we originally hypothesized. Note the second order fit as opposed to a linear fit. This is due to the parabolic nature of the movement requiring a parabolic change in grip force as they neared the apex of the movement when plotted in time. Except in three cases of the upward movement (**Figure 12**) the participant had a significant, but weak increase in force during the movement to the apex. In all cases of the downward movement however, the participant significantly decreased their force on the puck. The decrease in force had a much stronger correlation suggesting that the subject chose an initial lifting force close to what they needed and then slightly modified their grip during the lift. This may be why only some cases were significantly positively correlated on the upward movement as the participant could have initially overestimated their starting force and then decreased during the lift of kept their grip force the same. On the downward movement, in all cases the force was higher than needed when working with gravity so there was a large decrease in force.

**Figure 11:**
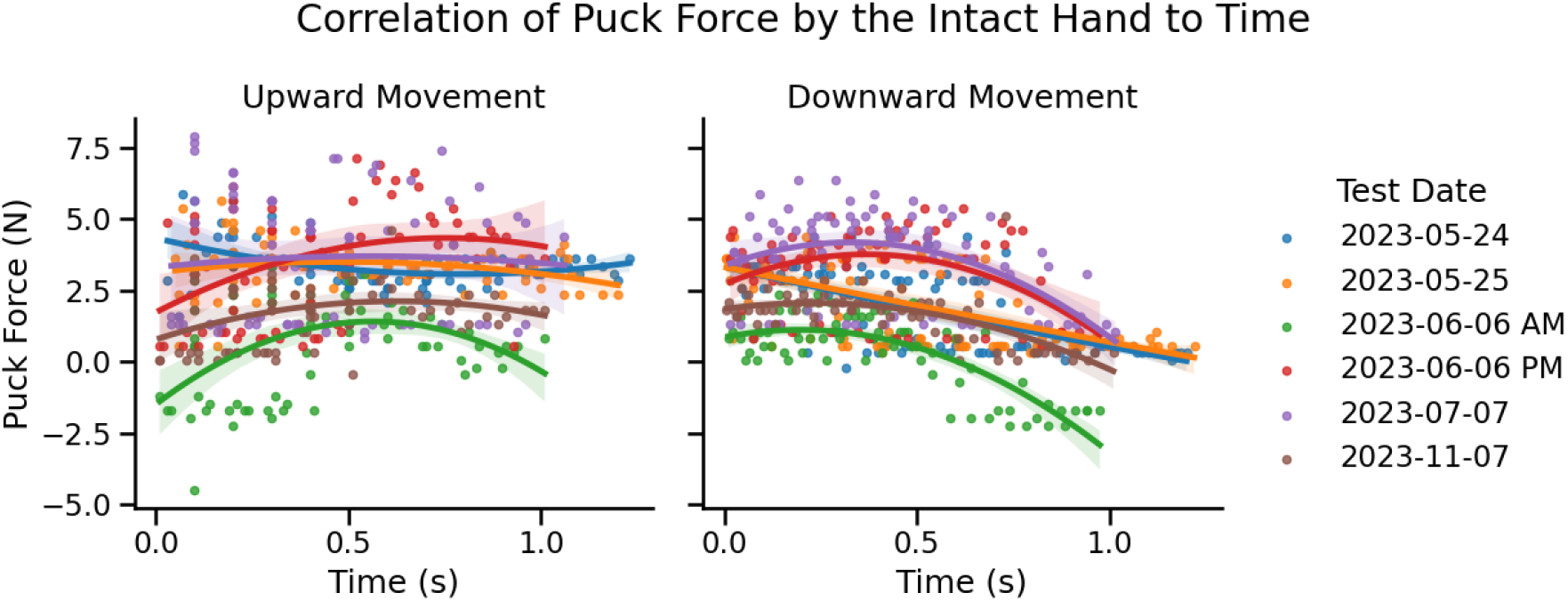
The correlation of the force from the participant’s intact hand with puck motion. On the upward movements, the participant’s intact hand showed a moderate correlation to the trial time and on the downward movements, there is a strong negative correlation. This indicates that the participant increased their grip force as they rose the puck from the table, and they decreased their force on the path towards the table.

**Figure 12:**
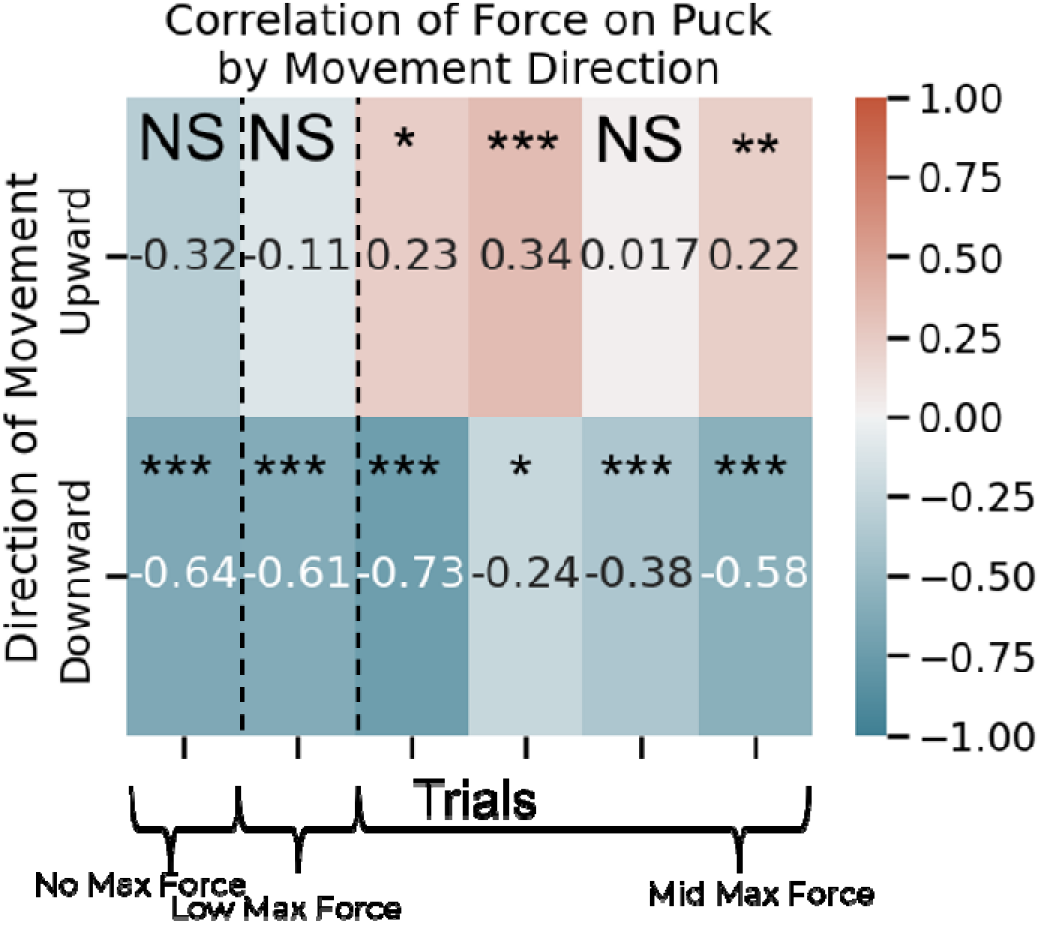
The correlation coefficients of the force from the participant’s intact hand on the puck during movement. Each column is a different trial, and the trials occurred in order from the left column to the right column. The forces applied by the participant’s intact hand correlate well with the direction of movement. In 3 out of 6 cases, the participant had a weak but significant correlation of increasing force as they rose the puck (redder), but in all cases of lowering the puck, the participant had a strongly significant tendency to reduce force (bluer).

The combined analysis of the force controlled prosthetic grip and intact hand force during motion shows that they both increase force during the upward movement, but only the intact hand appears to be reducing force during the downward movement. However, during testing we found that the TASKA hand could not make a fine enough grip change to lower the amount of force it has on an object without dropping an object. A single value grip position change was often enough to let the puck slip or fall from the hand.

### Events During the Trials

Usage of the force controller combined with stimulation generally resulted in more slip and drop events during the study despite equal numbers of force and velocity controller trials with and without stimulation (**Figure 13)**. Recorded events were labeled as slips, drops, pickups, steadys, stutters, pauses, table grasps, and shatters. Globally, these events can be categorized as errors (slips, drops, pickups, shatters) and uncertainty events (steadys, stutters, pauses, table grasps). The participant has time before the experiment to move the puck before testing to make sure they felt they could complete the task, understand the magnitude of the shatter force, and how the stimulation feels in comparison to the amount of force they are outputting when moving the puck.

**Figure 13:**
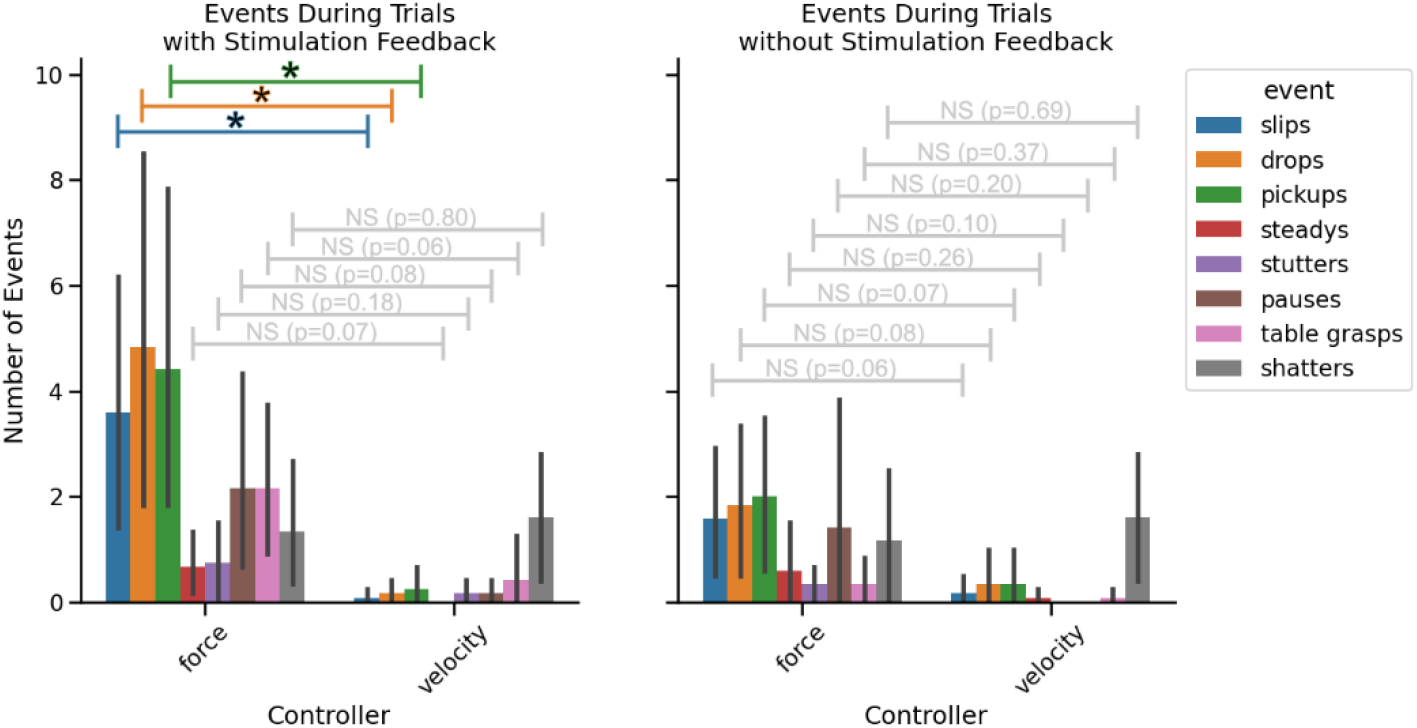
Error and uncertainty events during the puck movement task. The participant had many more marked events during use of the force controller, but notably, the number of shatters was the same with a force of velocity controller. This indicates confidence in the velocity controller but equal grip force modulation performance.

The error events in detail are as follows. A slip is an event where the puck moves within the grip but does not fall from the hand. A drop is any case where the puck falls from the hand during the trial, not including the instances at the end of the trial where the participant drops the puck back to the table. A pickup is each additional time during a trial the participant picked up the puck after a drop with their prosthesis. Note that the number of drops and pickups are not necessarily the same as the participant sometimes caught the puck with their intact hand or, very rarely, with their prosthesis. Finally, a shatter occurs when the participant uses a force larger than the software defined shatter force. The participant was notified of a shatter by a sound from the testing setup.

The uncertainty events recorded instances where the participant needed to pause to make sure they would not cause an error. Steadys were instances where the participant used their intact hand to steady the puck in their prosthesis to either check their grip or readjust grip. Stutters were instances where the subject moved in a jerky movement. This indicated that the subject thought they were about to drop the puck, and they reacted as if they needed to catch the puck. A pause occurred when the participant stopped in the air before continuing. A table grasp were additional times the participant regrasped the puck at the beginning of a movement or at a pickup error event before lifting the puck off the table successfully.

**Figure 13** shows the usage of a force controller with stimulation resulted in significantly more error events compared to the velocity controller with stimulation, except in the case of shatters. The uncertainty indicators with the force controller trended towards significance, but did not reach significance. In the case without stimulation, the force and velocity controllers did not differ significantly in their number of events.

The main result from **Figure 13** to note is the introduction of stimulation seeming to increase the number of error events that lead to the puck slipping or dropping from the prosthesis. Despite this, the number of shatters were not significantly higher with the force controller. This indicates that stimulation led to an under estimation of the required force applied to the puck during movement or the intention to reduce force on the puck. Over estimations causing shatters were just as common with the velocity controller or without stimulation as there were no significant differences with between the shatter cases when compared across all conditions.

If the participant were attempting to reduce their grip force, one may ask why there was no significant negative correlation of their grip force to the puck movement. This is due to the puck movement data only encompassing successful trials when the movement was completed. This eliminates any trials where the participant dropped the puck from loosening their grip.

Combined with the results in **Figure 11** that show the participant’s intact hand has a strong relationship with lowering their grip force during end of a movement, the addition of stimulation may be leading to an intended decrease in force when moving with gravity for this specific task. However, this intention resulted in the release of the puck instead of the reduction in force by the device due to the prosthetic hand’s design.

### Grasp Intent Contained in EMG

To investigate if the participant intended to reduce their grip force during the trials, we measured the correlation of the participant’s EMG with the puck displacement. We hypothesized that the mean absolute value of the participant’s EMG would negatively correlate with the puck’ movement. In the recordings consisted of four flexors (the Pronator, FCR, FDS, FCU) and four extensors (the Supinator, ECRB, EDC, ECU) the flexor muscles have a mean above zero while the extensors generally have no activity until they are needed to open the hand (**Figure 14**).

**Figure 14:**
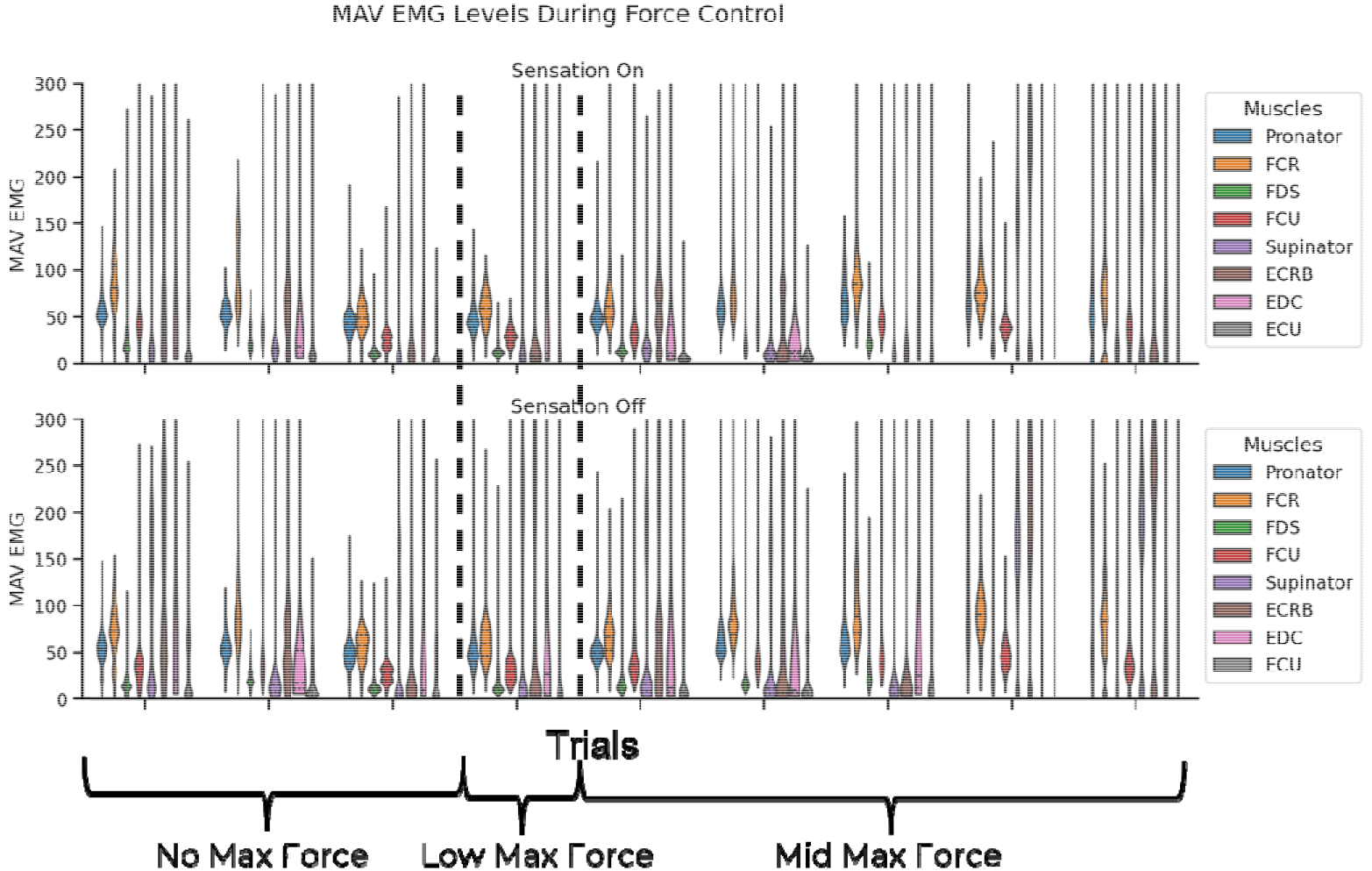
Flexor and extensor EMG distributions during the puck movement task. Each column is a different trial, and the trials occurred in order from the left column to the right column. The EMG activity of all muscles during the force control tests. In general, the flexors had higher activity and centered on a mean value above 0. The extensors were used mostly to open the hand, so their force did not need to be as high as the flexors.

Figure 15 shows the correlation of the mean absolute value of the participant’s EMG to the puck’s vertical displacement with the force controller and stimulation. The figure shows that with a high shatter force (the first three trial dates) the flexors muscles in the first four columns have a generally significant negative correlation on both the upward and downward motions.

**Figure 15:**
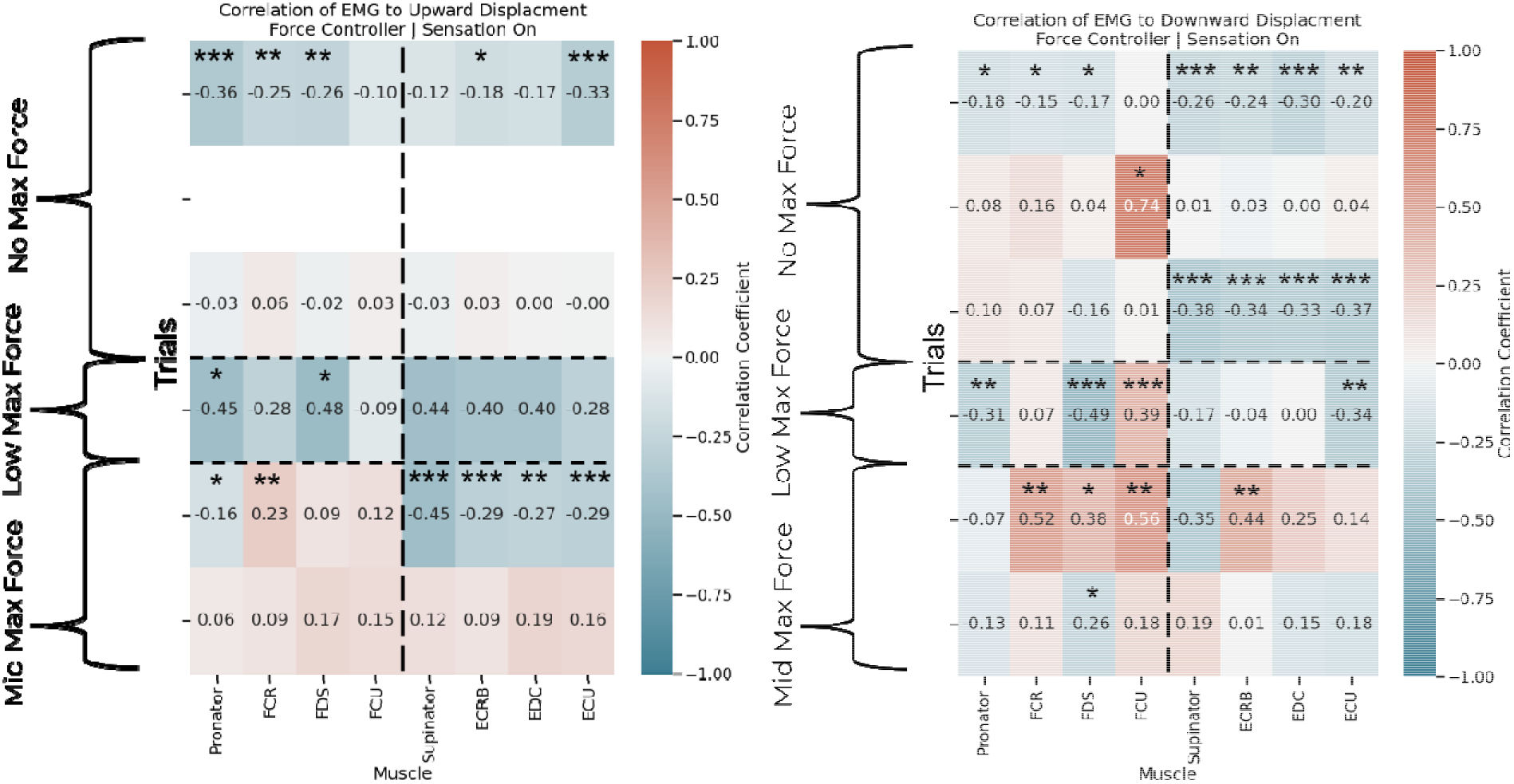
The correlation of the participant’s EMG with the force controller and stimulation to puck movement. Each row is a different trial, and the trials occurred in order from the top row to the bottom row. Before adding a shatter force on the fourth trial date, the participant’s EMG has a negative trend (bluer) with the puck movement indicating a general intent to reduce force. On the fourth trial date, the upward section of the flexor EMG showed some significant reduction in force towards the apex, but the downward section shows no significant decrease in EMG. After the fourth trial date, the EMG is not significantly different from zero.

This indicates that with a shatter force that was too high, the participant was generally always trying to reduce their grip force likely from starting at a high initial force. On the fourth trial date where the shatter force was the lowest, the upward tracking generally shows a decrease in EMG amplitude, but the downward section of the EMG shows a no significant change in the EMG. After the fifth trial date, when the shatter force was increased to a moderate level the participant still found difficult the downward section of the flexor EMG is not significantly different from zero indicating an intent to keep EMG at a constant level. Also note that on June 6^th^, the extensor EMG was significantly negatively correlated on the upward section on the EMG. This indicated it was opposing the flexors less on the rise of the puck.

In Figure 16 the early days before the addition of a lower shatter force again show a general desire to reduce force. This is shown in the significant negative correlation in the flexors and some positive correlation in the extensors. During the fourth trial, the day with the lowest shatter force, the flexor and extensor EMG all show a significant positive correlation with the upward movement suggesting the participant was co-contracting their muscles harder as they lifted the puck to hold the puck stable. The positive correlation of the flexors and two flexors on the downward section suggests they kept this co-contracting during the downward movement. Compare this to the same case with stimulated feedback where the participant was still trying to reduce force. After the fourth trial, one upward trace showed a significantly negative correlation of the EMG with displacement, but both downward traces show significant reduction in EMG suggesting they were intending to reduce grip force during these trials.

**Figure 16:**
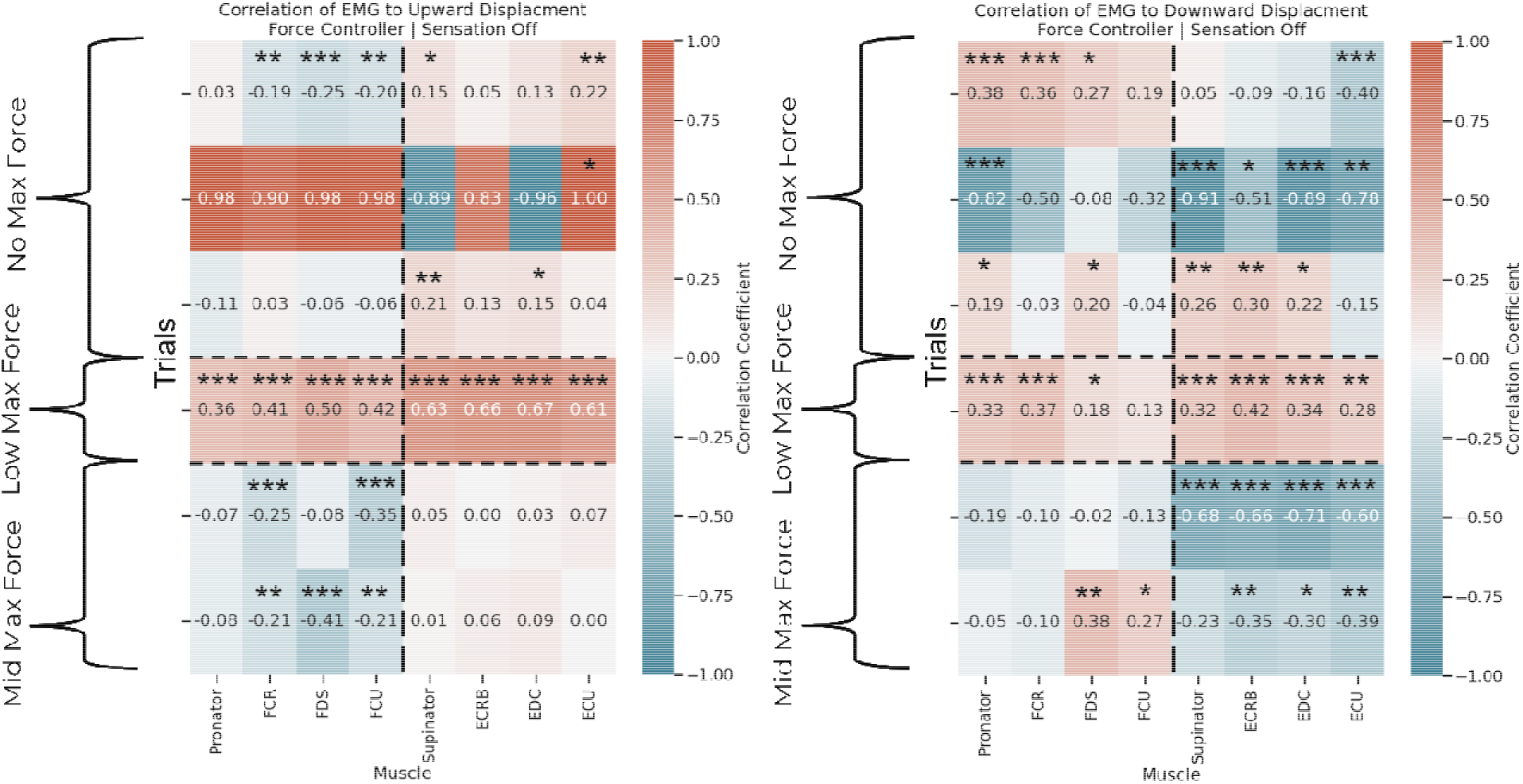
The correlation of the participant’s EMG with the force controller and without stimulation to puck movement. Each row is a different trial, and the trials occurred in order from the top row to the bottom row. Before adding a shatter force during the fourth trial date, the participant’s flexor EMG has a negative trend (bluer) with the puck movement indicating a general intent to reduce force. On the fourth trial date, the upward section of the EMG shows the participant was significantly increasing force in all muscles indicating an increasing contraction during the test and a constant contraction on the downward section of the movement. After the fourth trial date, the EMG is not significantly different from zero.

Figure 17 shows the correlation of the participant’s EMG with their existing velocity controller with the addition of stimulation. With a high shatter force threshold, the participant’s flexor EMG shows no significant trend on the upward movement, but the extensor muscles tend to have a significant positive correlation on the downward movement. This could indicate the participant intended to loosen their grip on the downward section to their movement, but the participant also stated they “pre-load” their muscles before movements. By this, he means he starts a contraction early that is below the threshold to activate his velocity controller so that when it is time to move the controller can be activated quickly. He may have done this to quickly open after setting the puck down. After the fourth trail date, with the moderate shatter force, the participant shows signs of co-contracting their muscles as seen in the generally positive and significant correlations of EMG to puck movement on the upward sections of movement. However, the downward sections of movement show both a significant reduction in EMG and increasing in EMG. The reason for the co-contractions in the upward movement may be due to a shift in strategy the participant stated he had. He stated that when he wants to make sure he doesn’t drop an object that is important, like a more fragile puck in this case, he starts contracting to make sure he doesn’t drop it. He contracts at a level that is below the threshold to activate his prosthetic controller but does this anyway to feel this the object is more secure. The different positive and negative correlations on the downward movements may then be a relaxation of this co-contractions or a continuation of them.

**Figure 17:**
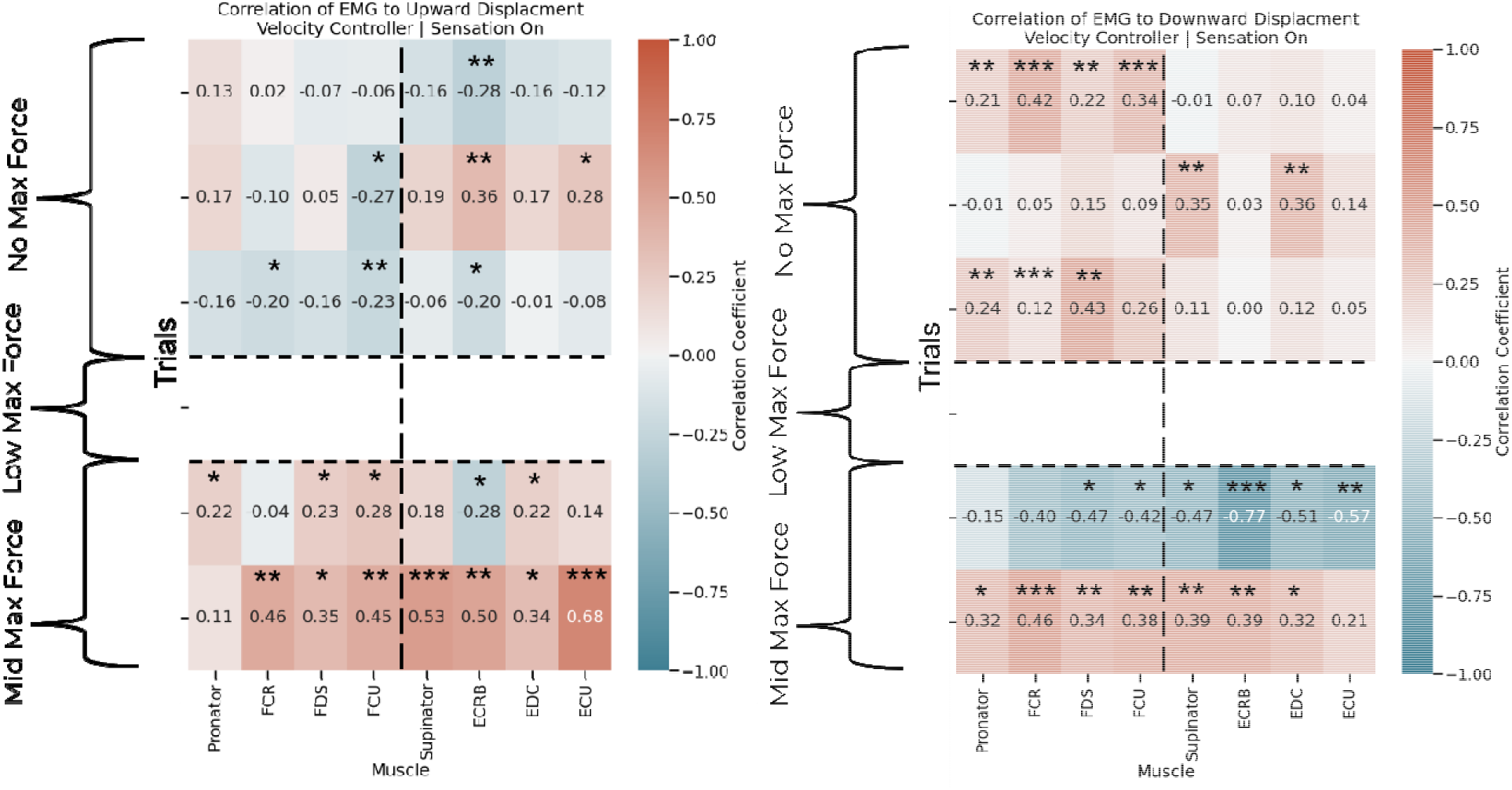
The correlation of the participant’s EMG with the velocity controller and with stimulation to puck movement. Each row is a different trial, and the trials occurred in order from the top row to the bottom row. Before adding a shatter force during the fourth trial date, the participant’s flexor EMG had no significant correlation with the puck movement, but the extensors generally were positively correlated with puck movement on the downward movement. This could indicate an intent to release the puck, but the participant also states that he “pre-loads” his movement before doing them so he also could be starting the opening contraction with his extensors early. The last two trial dates with a moderate shatter force show a generally positive correlation of all EMG to puck movement on the upward movement, but a significantly negative correlation and positive correlation on two different days for the downward movement showing a mixed intent.

Figure 18 shows the correlation of the participant’s EMG using their velocity controller without stimulation. This is the typical way the participant uses their device at home. During the first three trials, the period with a high shatter force, the participant generally relaxed all of their muscles, shown by a significantly negative correlation on the upward movement and no significant correlation on the downward movement. This is consistent with our expectations of how a participant usually use a velocity controller. The velocity controller allows the participant to grab the puck at the start of a movement and then stop contracting when moving the puck if the force was not too high or too low. This is one of the main benefits of a velocity controller as it saves battery life. If there is enough friction between the prosthesis fingertips and the puck, there is no need to adjust the grip. During the third trial, the opposite trend started happening, The participant’s flexor EMG during the upward significantly increased and then they decreased their extensor EMG. This indicated an attempt to keep an increasingly higher force. This would not fail the test at the shatter force was too high to reach this day. After the third trial date, on the days with a moderate shatter force, the participant had no significant trend in the upward movement, but a general co-contraction on one day of the downward movement. Notably, these correlations were especially high. This day also aligns with the day the participant constantly increased their force with their velocity controller with stimulation. They may have done this again to “feel” like they were holding the puck harder, but this level of EMG did not move their hand.

**Figure 18:**
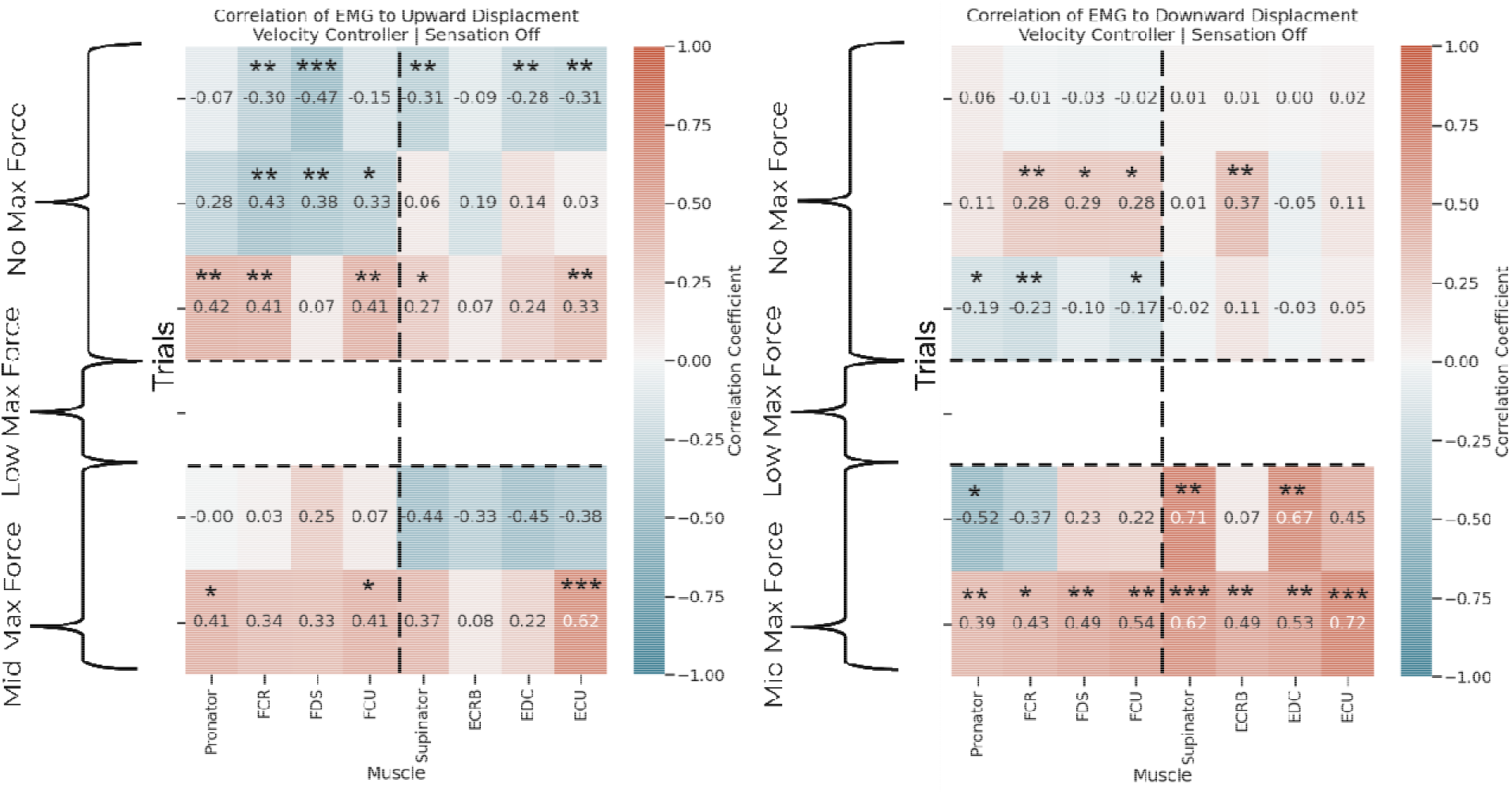
The correlation of the participant’s EMG with the velocity controller and without stimulation to puck movement. Eac row is a different trial, and the trials occurred in order from the top row to the bottom row. Before adding a low shatter force on the fourth trial date, the participant’s flexor and extensor EMG had a significantly negative correlation with the puck movement, but no correlation on the downward movement. On the third trial date and onwards the participant’s EMG tended to have a positive correlation that was significant across all muscles.

There were differences between the muscles’ signals used by the force controller versus the participant’s velocity controller. The force controller uses the implanted EMG electrodes so the EMG recordings we have are exactly what are used by the controller. The velocity controller uses the electrodes in the participant’s socket instead and they record the surface activity of the wrist flexor and extensor muscles as well as any cross talk from nearby muscles. For this reason, the EMG recordings we have for the velocity controller are not exactly what the controller has as an input. In fact, looking at our data, the velocity controller EMG amplitudes do not align with when force was released from the puck (Figure 19). Common in all tests was the appearance of a large spike in the extensor EMG signals that stopped before force was released from the puck. It is possible our electrodes are missing other muscles that are part of the surface recording that form the surface signal. However, the muscles we recorded from are the most important muscles in grip and object manipulation and should have contained any automatic intent to modulate grip.

**Figure 19:**
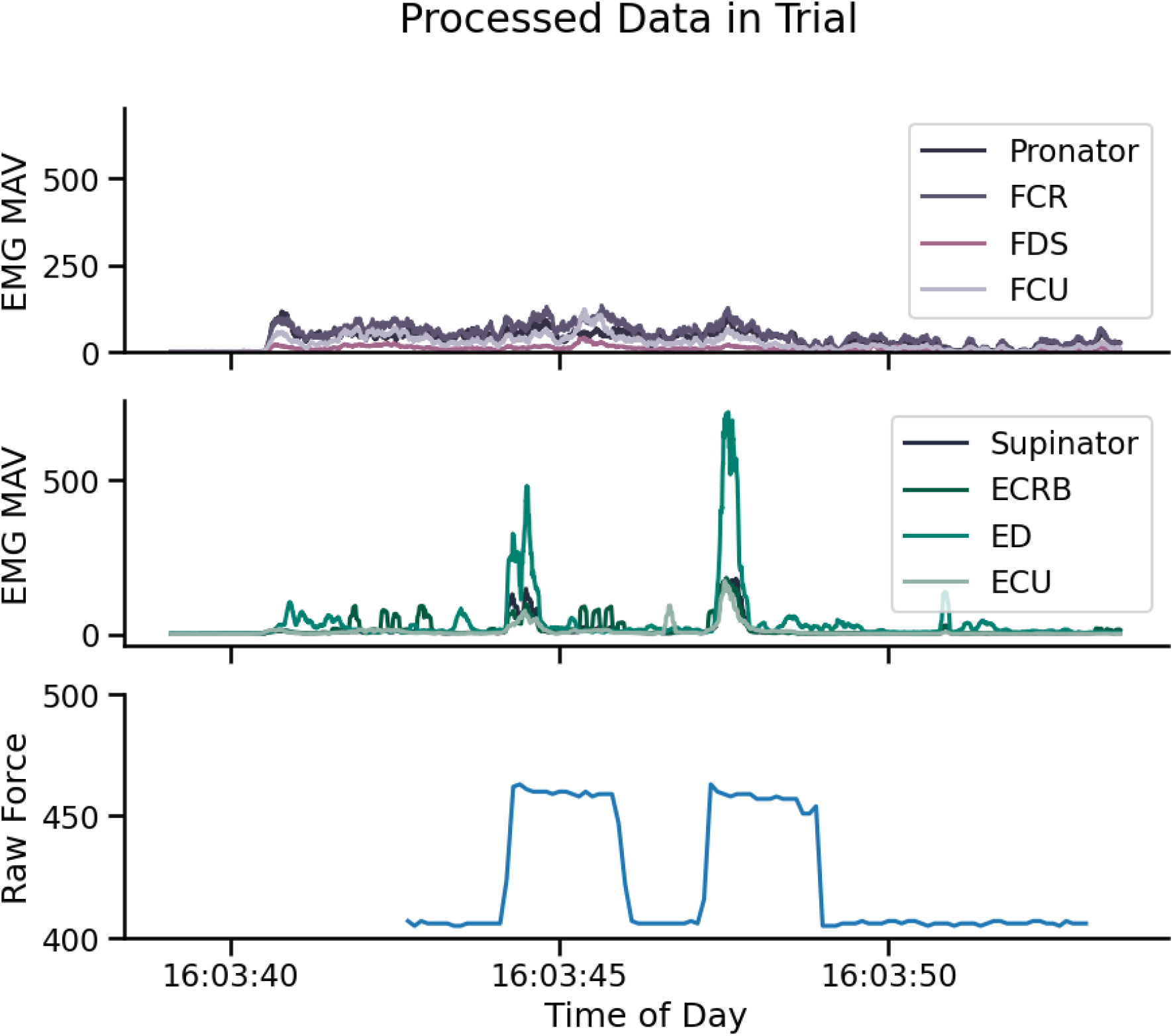
An example of the velocity controller behavior not matching with the recorded EMG. In this example raw recording of a trial with the velocity controller, extenso EMG traces start and end earlier than expected suggesting that the implanted EMG electrodes to not get the same signal at the electrodes in the participant’s socket.

When all of the EMG analysis is taken into account, there are general conclusions for how the participant used each controller. With stimulation, the participant generally reduced their force especially at the high shatter force thresholds. At the moderate threshold, they kept their EMG constant. Without stimulation, the participant was generally always reducing their force except at the low shatter force. In this case, without stimulated feedback, they were slowly increasing force at all times to make sure they did not drop the puck. For the velocity controllers, the participant’s EMG at the high shatter forces shows they generally initially grab the puck and stop contracting. At the lower shatter thresholds, there appears to be a shift in mindset where the participant starts co-contracting to feel like they are holding the puck regardless of their stimulated feedback. This level of EMG however does not activate their controller to move. It does however give evidence that the participant wants to use their EMG as a sense of their force on the object.

### Distance from the Barrier

The final analysis of the study showed the distance the participant moved the puck above the barrier depended on the controller and if they were given stimulation (Figure 20). The highest above the barrier involved movements using the force controller without stimulation. Next was force control with stimulation, then velocity control with and without stimulation respectively. The lowest is when using the intact hand and was not significantly different from when using stimulation with the velocity controller.

**Figure 20:**
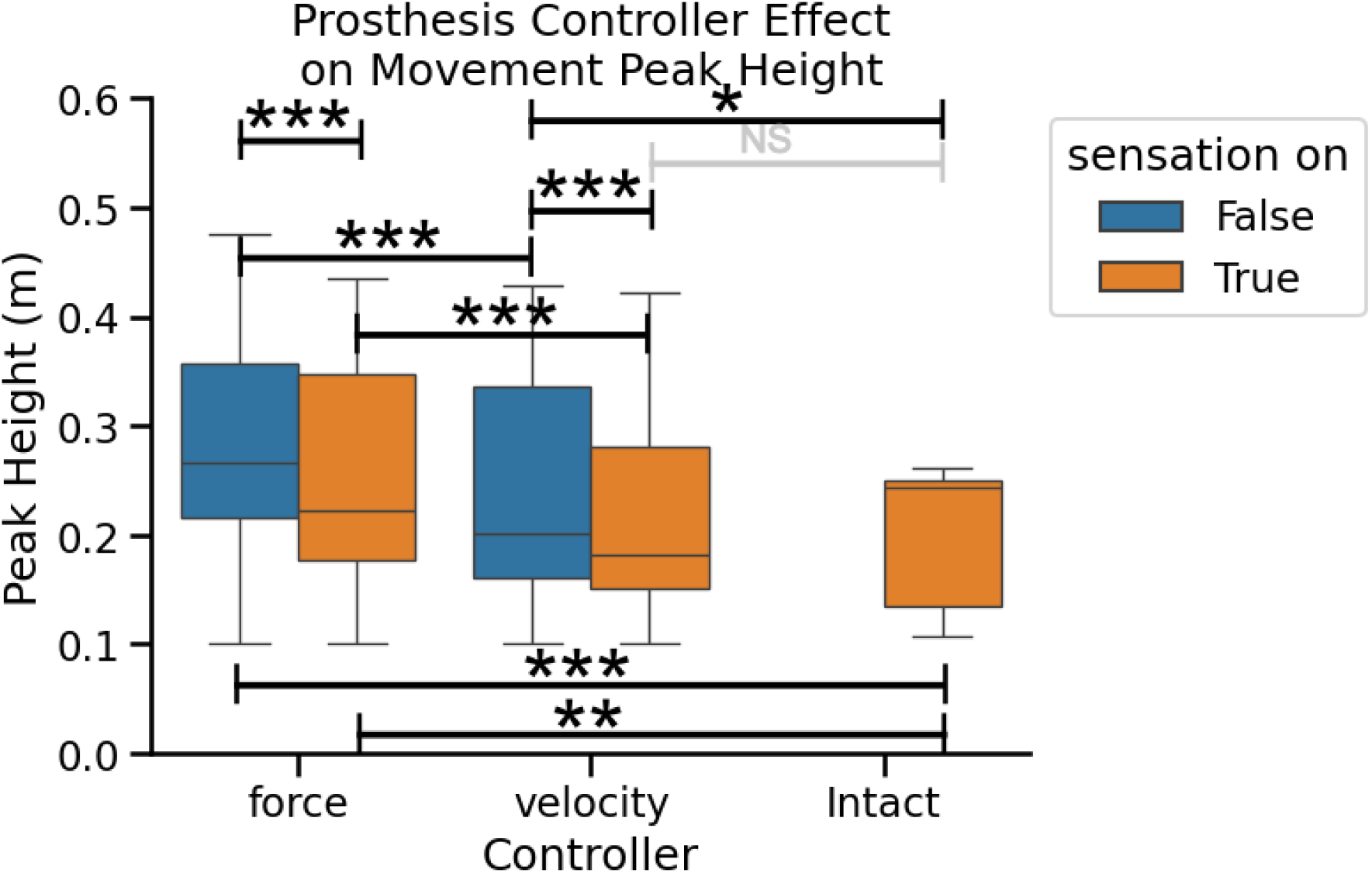
The difference in the apex of the participant’s movement of the puck with the force and velocity controller with and without stimulation. The height the participant moved the puck above the barrier depended on both what controller they used and is they received tactile feedback from stimulation. The participant using the force controller led to the highest movements, the velocity controller had lower movements, and the intact hand had the lowest movements. The application of stimulation lowered the movement heights with either controller significantly and lowered the height of the velocity controller with stimulation enough not to be significantly different from the intact hand.

One theory for why the participant moves the puck higher with the force controller is due to a reduced confidence with the force controller. However, referring back to the uncertainty metrics from Figure 13, there was a trend towards more uncertainty with the force controller, but it was not significant compared to the velocity controller. The addition of stimulation as a source of tactile feedback lowered the movements nearer to the barrier for both controllers, perhaps indicating a better understanding of where the puck is in space. The velocity controller’s movements may differ from the force controller movements because the subject may have better understanding of the velocity controller as this is the controller he uses every day at home.

## Discussion

### Overview

The initial goal of this study was to show evidence that the addition of stimulated tactile feedback to a basic object movement task would lead to changes in grip force during the movement. Our study was modeled after similar work with able-bodied individuals which found the intact tactile system integrates with the motor system at such a fundamental level that our hands correct for load force changes automatically (Cole and Abbs, 1988; Johansson and Westling, 1988; Flanagan and Wing, 1993). This integration happens at such a fast speed and at such a basic level that it likely occurs in the brain stem (Scott, 2016; Loutit and Potas, 2020). As a result, we hypothesized that artificial tactile feedback supplied during an object manipulation task would also lead to grip force changes because the brain stem is thought to be robust to noisy signals or malformed tactile feedback (Loutit and Potas, 2020; Fujita, 2021). The results of this study show that when given direct control of force, participants increase their force on objects when they lift them. They also may intend to reduce their force when they set object down with a force controller, but current commercial prosthetic devices are not capable of reducing grip force without first opening. Artificial tactile feedback seems to enhance this intent to reduce force as seen in the increase of slip and drop events when stimulation was added to the force controller and stimulation appears to enhance the participant’s internal perception of their body. One of the main factors that needs to be addressed if a force modulation to be possible with a prosthesis is its design.

Modern myoelectric prostheses are designed to convert the electrical signals from the muscles to a velocity of movement and get to a set position for a static grasp fast. Unless an object is particularly fragile, there is likely no need to have a high resolution of grip positions for a prosthesis, and in those cases, a prosthesis user would likely use their intact hand, if they have one, to care objects they particularly care about. The benefits of this design are a low battery drain due to infrequent need to activate the motors and a low effort on the part of the user when carrying a rigid object as the user only needs to activate their muscles to set a grip and ideally, they could stop attending to the object during transit. However, many objects do not fit in this ideal scenario. Using the intact hand for important tasks will lead to an over reliance on the intact hand and could lead to overuse injuries. In fact, almost twice as many individuals with one upper limb missing report musculoskeletal issues compared to intact individuals and is hypothesized to be from an overuse of the intact limb even in tasks that do not require the intact hand (Chadwell et al., 2018).

We hypothesize that bringing prosthesis use closer to the intact experience will lead to a reduction in this intact to prosthesis use mismatch, but it will require large changes to how prostheses are designed. Mainly, this may involve the shift from a velocity controller to a force controller. Our muscles output a force, not a velocity. Direct control of output force may be preferable to individuals with limb differences despite the larger energy draw on their device and muscles. We see this with many people’s preference for body powered prostheses due to the direct force feedback they feel (Brown et al., 2017). The intact hand holds an object with some constant or changing active muscle contraction, so it stands to reason that a more intuitive control scheme would involve a direct control of force by some constant muscle contraction. We saw this behavior in this study where the introduction of a shatter force led the participant to feel the need to hold a contraction with his velocity controller, even though it does not activate any movement by the controller. He stated he even does this at home when carrying objects, he does not want to drop, just to “feel” like he is holding the object.

In addition to direct control of force, there may need to be a shift to backdrivable prostheses, or devices that can change force on an object within a static grip position. As we found in this study, the participant showed the intent to change their grip force given a force controller, but the coarse resolution of the device in the study did not have the ability to change the force on the object without first dropping it. This is common in modern prosthetic devices because the need is to get a good enough position as fast as possible. However, even “good enough” still led to no statistical difference in over corrections of positions with the velocity controller compared to the force controller as seen in Figure 13. Both cases would be helped by a backdrivable device that could reduce force.

### Force Modulation During Movement

During the movement of the puck over a barrier, there was a significant correlation between grip force and the upward movement of the puck (Figure 9), but no significant correlation on the downward motion when using a force controller (Figure 10). The velocity controller showed no significant correlation between grip force and puck movement. These results indicate that when the participant has direct control over force, they intuitively increase their grip force in opposition to gravity and the inertial force of the puck, but they keep a constant force from the movement’s apex until the puck is set on the table. The participant’s intact hand shows the same increase in grip force but also shows a decrease in grip on the downward motion (Figure 11 and Figure 12). The participant’s lack of a decrease in force with the force controller is likely more to do with the prosthesis’s inability to reduce force and less to do with a lack of intent to reduce force.

There was no difference between using the force controller with and without stimulation for this analysis which indicates that in this context, just the control of force and the experience of moving the puck was enough to create the desire to increase force. This increase is unlikely to be a conscious decision as it unlikely that the participant would decide to increase grip as the puck moved vertically instead of picking a force at the start of the test.

The lack of a significant correlation between grip force and movement with the velocity controller was expected as the typical usage of these controllers consists of picking a grip position at the start of a movement and then not changing the grip during the movement. The only reason a velocity controller user would change their grip during the motion would be if the object were slipping, but Figure 13 shows slip events were extremely uncommon with the velocity controller.

### Prosthesis Output Force Resolution

During all trials with a prosthetic hand, the force on the puck plateaued during movement. The main reason for this, as described above, is due to the prosthesis not being able to reduce force without first opening. Our validation data (**Supplementary Figure 1, Supplementary Figure 2, Supplementary Figure 3**) shows that the device can closely follow the macro levels force changes, but the jitter in those force traces is due to the hand slightly closing more or opening more than it needed to. The validation data was also taken on an object that was sitting on a table so there was no chance that the hand opening too much would cause the object to slip or drop. While large changes in force were possible, our results imply minor changes were not. The high number of slips and drops with the force controller show the participant’s intent to modulate their grip resulted in the hand opening too much, but equal “shatters” among all controller cases which indicates that their initial acquisition of the puck was identical.

Note the velocity controller can make smaller changes in grip than the force controller. This is because the force controller is limited by a small number of positions between 0 and 255 while the velocity controller sends a voltage through the socket to power the motors directly. A straightforward solution to giving the force controller the same possible resolution as the velocity controller is use the socket connection to the device and created a feedback system for force to voltage instead of grip position. The prosthesis in this study also for control of the motor torque through the same connection as the current force controller. We did not pursue either option in this study to not damage the participant’s personal socket in the former case and not damage the prosthesis by applying a high motor torque for too long in the latter case. However, with the limitations of the current force controller design evident in this study, there is merit to creating an in lab socket we can modify or carefully designing a force controller of motor torque that minimizes damage to the hand.

### Events During the Trials

Despite no differences in the correlation results between the force controller with and without stimulation, the force controller with stimulation had more slip and drop events during the trials compared to any other condition (Figure 13). The higher number of slips and drops indicate that given the addition of stimulation, the participant tended to reduce their force during the trial which led to the prosthesis opening too much. In our control results using the participant’s intact hand, we saw a significantly strong correlation between grip force and the downward motion of the puck (Figure 11 and Figure 12). It is possible that the addition of stimulation to the force controller enhanced the intent to reduce force on the puck, but the prosthesis overcorrected and opened instead.

Interestingly, shatters were the only error event that had no significant difference between controller conditions or stimulation conditions. This result implies that the regardless of if the participant was in control of the force or the velocity, they overcorrected their force in the same proportion of times between conditions. For the force controller this is caused by too strong of a contraction in a direct over estimation of grip force at the start or during the trial, but for the velocity controller, a shatter occurs from an over estimation of the initial grip position needed to create a force. Before all tests, the participant had the opportunity to move the puck to learn the force they needed to apply and yet these over corrections occurred in the same proportion at some point in the trial. The implication of this result is that direct control of force and the addition of tactile feedback is not enough to prevent an overestimation of required force. A future solution to this may be some proprioceptive signal that allows the participant to feel how closed their hand is in addition to their vision like an able bodied individual has. The stimulation in this study only activates when the subject comes into contact with the puck and by then it may be too late to correct their grip size, but a proprioceptive signal may tap into the existing sensorimotor control loops to estimate grip. At the time of writing, the addition of proprioceptive stimulation is an active area of study.

Among the other events, there was no statistical difference in the events that indicate uncertainty (steadys, stutters, pauses, table grasps), though the values suggest given more data, the divide between uncertainty in the force controller and the velocity controller could become significant. We hypothesize this would happen due to the participant stating he found the force controller frustrating and error prone during the test due to the number of drops. He also stated that he found it took more effort to keep contracting his muscles to prevent an object from falling from his hand when he knows if he stopped contracting with his velocity controller, an object would not slip out. However, as we see in the EMG data with the velocity controller and of the participant’s own admission, he actually does keep contracting his muscles with a velocity controller when he does not want to drop an object, it just doesn’t activate the controller.

Other than improvements to a prothesis, this effort the participant experienced with the force controller should be addressed in the future. The current iteration of the force controller did not account for any passive elements of the hand in keeping the hand within a grip, however, there is an inherent stiffness to grip which should have been added to the controller to reduce the active effort of the participant (Ambike et al., 2014).

### Grasp Intent Contained in EMG

The initial results showing an increase in grip force to the apex puck movement but no decrease (Figure 9 and Figure 10) seemed to imply that force control with a prosthesis differed from the increase then decrease in force seen with intact individuals (Cole and Abbs, 1988; Johansson and Westling, 1988; Flanagan and Wing, 1993). However, the combination of force control and stimulation showed a significantly higher slip and drop rate which led to the hypothesis that the participant had the intent to reduce grip force that was not being correctly output by the prosthesis. This behavior would be consistent with the behavior of the intact hand during the trial which showed a strong correlation with lowering grip force during the trial movements (Figure 11 and Figure 12).

When using a force controller, the correlation between the mean absolute value of EMG and puck movement shows a significant negative correlation that suggested the intent to reduce force during the trial. This negative correlation was stronger before the introduction of a shatter force (Figure 15 and Figure 16). A possible reason for a stronger negative correlation before the addition of the shatter force is due to the participant having the ability to use as high a force as they want to lift the puck which they then intend to reduce after the apex of the movement. After the addition of the low shatter force, the participant kept a more constant contractile force with stimulation, but generally increased their EMG without stimulation towards the shatter threshold. This seems to indicate that the participant had a better idea of where the shatter threshold was with stimulation but was slowly increasing force without stimulation because the participant did not know where the shatter threshold was. With a moderate shatter threshold there was no significant correlation of EMG to puck movement with stimulation, but a general decrease in EMG when the controller was used without stimulation on the downward motion. This shows that the participant felt internally that the needed to reduce force or that they felt they were getting close to the shatter threshold when they hand no stimulated feedback.

When using a velocity controller, the participant’s muscles had a mix of correlations both positive and negative during the test (Figure 17 and Figure 18). More strong trends appeared after the introduction of a shatter force with one day showing a significant positive correlation and other with a significant negative correlation. Before the introduction of a shatter force the results indicated no significant intent to change grip force during the test. After the introduction of a shatter force, the participant stated that the had been holding a low level contraction during the test to feel like he was holding on to the puck. This is likely the source of the correlation with the movement. The desire to hold a contraction lends evidence to the idea that prosthesis users may find force controllers to be more intuitive.

Note however that the muscles that controlled the velocity controller were not the same as the muscles we recorded from. The velocity controller used electrodes over the muscles that flex and extend the wrist while we recorded from four flexor and four extensors muscles which are some of the main contributors to hand position. There would be overlap in the internal recordings we have, and the surface recordings used in the socket, but as Figure 19 shows, the muscle activity we recorded does not always align with the prosthetic hand’s behavior.

It is important to note that during these tests, stimulation changed with changes of force. The subject appeared to have the ability to increase force, shown by the positive correlation of force to puck displacement on the upward movement (Figure 9) but did not change when the subject intended to decrease force as shown by the non-significant correlation of force to puck displacement on the downward motion (Figure 10) compared with the EMG results showing the participant trying to reduce grip force. This means that the feedback the participant received did not match their grip intent. Increasing the grip resolution would improve the output force of the device, but also amplifying even minor changes make these minor grip changes distinct in the sensorimotor system. Even without changes to the device some component of stimulation could be based on the EMG level of the participant’s muscles. Note that the participant may not need to perceive the changes. The aim is for the brain stem to receive changing feedback in reference to the participant’s motor output. Another solution is to use the movement of the puck in a model of the inertial forces from the puck to the hand and supply artificial changes in feedback.

### Distance from the Barrier

This study shows a two-part trend in how the participant moves the puck relative to the barrier in the test. The participant moved the puck statistically higher when using the force controller compared to when using the velocity controller and moved the puck higher with either controller when moving the puck without sensation. In comparison, the participant moved the puck the lowest with the intact hand and the combination of velocity control with stimulation produced a movement height that was not statistically different from the intact hand’s movement.

The distance the participant moved the puck from the barrier provides a measure of the margin of safety the participant wants to keep when preventing the puck from striking the barrier. Note that in no case did the participant ever strike the barrier and they were instructed to move the puck in whatever way they wanted when transferring from one position to another. Moving the puck higher would lead to a larger path length for the puck and extend the trial time. Comparing the results of this safety margin to the uncertainty events in Figure 13, the latter results indicate no significant conscious uncertainty in the participant’s ability to moves the puck in space, but the former results show an inherent uncertainty that the participant may not be consciously aware of. This uncertainty seems to be significantly reduced given the presence of tactile feedback.

One reason the participant moved the puck higher with the force controller compared to the velocity controller may be from their lack of experience with the force controller. The participant has used their velocity controller constantly at home for over a decade, but only used the force controller during the test. This would give them less than a day of cumulative experience. Regardless of controller, stimulation immediately reduced the height of the movement indicating instantaneous improvements in the path the participant took when moving the puck. These instantaneous improvements are consistent with other studies that show stimulation can immediately improve task performance for spinal cord injury participants (Flesher et al., 2021).

Another explanation for the participant moving the puck lower given stimulation may be from an update to the internal model of the body. All forms of sensory feedback constantly update our internal model. This has been shown to occur even with audio signals substituted for touch with able-bodied participants (Shehata et al., 2018) and with continuous, supplementary vibrational feedback for grip force modulation with participants with limb differences using prosthetic hands. (Cappello et al., 2020). In fact, Cappello found that supplementary feedback was only needed in the dynamic phase of grip to update the internal model of the body, not the static portion of grip (Cappello et al., 2020). Flesher showed functional improvements with touch are instantaneous (Flesher et al., 2021) and Risso shows touch optimally located on the internal body model easily integrates with the other senses even without any patterning (Risso et al., 2019). It is possible that just getting touch on the hand that aligns with where the participant sees their hand touching an object and having to actively use the muscles is enough to update the body model with there the hand is in space. In previous our studies we have seen the addition of stimulation can change the perceived phantom limb length which shows stimulation does update the internal mode (Cuberovic et al., 2019).

### The Importance of Slip

This study focused on supply grip information purely through pressure, but the shearing of skin during slip is also an important part of grip force modulation. Previous studies show the sensation of slip on the fingertips is important for instantaneous grip force modulation. Even perturbations of the hand that don’t elicit shearing of the skin don’t induce grip changes (Cole and Abbs, 1988). Other studies show that humans and animals have evolved different structures just to understand slip alone (Schwarz, 2016). Humans developed ridges on their fingertips so well-tuned to detecting slip that single ridges of a fingerprint will start to signal local slips well before any global slip has occurred (Schwarz, 2016). Slip is important for not dropping objects and countering perturbations. Pressure information is also important in quantifying the current force on an object, but the slip force gives information on how other forces perpendicular to the fingertip like gravity are affecting grip. During the movements in this study, the puck was held by the participant vertically. This position in the hand should produce a shear force on the fingertips of the prosthesis that was not recorded or used in the stimulation. The puck horizontal could have been horizontal so the force of gravity pointed towards the faces of the sensorized puck, and that changing force was converted to stimulation, but the movement side to side that would cause a shearing force would not be captured. Both pressure and slip are necessary to fully capture the feedback required for grip force modulation.

Other than introducing shear sensors into the study, an alternative solution to estimating slip forces is to model the forces parallel to fingertips based on the motion capture data and add this factor to the stimulation. The difficult aspect of this solution or the addition of shear sensors is how to give a signal that is feels or is used like slip. It is however possible the brain stem can also use this information without the perceptual qualities to slip if given in the right context.

### Intrinsic vs Extrinsic Muscles

This study recorded from the extrinsic muscles of the hand alone because they are the major muscles that control grip and because they are traditionally used when controlling a prosthetic device. This is the same rationale used when choosing which muscles to implant in the participant. However, investigations into the muscles used in grip have found the extrinsic muscles control the coarse level of grip and help maintain a certain hand shape while the intrinsic muscles rotate the fingers into place and control for the fine changes of grip by modulating the force on each finger during object manipulation (Long et al., 1970; Maier and Hepp-Reymond, 1995). Obvious intrinsic muscles that contribute to grip are the muscles of the thumb, without which opposable grip is not possible, but even the intrinsics of the rest of the fingers are constantly changing their forces as an object moves in space and within the hand.

This is most important during low forces. Intrinsic muscles have a high correlation with grip force at low forces and the intrinsic muscles could have a primary role in force modulation at low forces (Maier and Hepp-Reymond, 1995). The pattern of extrinsic activity differs highly with if a person is actively griping an object or not. For example, the Flexor Digitorum Superficialis (FDS) does not change its contractile activity with force unless there is an object in the hand (Long et al., 1970).

Intrinsic and extrinsic muscles act together during object manipulation and without the existence of intrinsic muscles with a participant lacking a hand, recording the true intent to modulate grip may be difficult. It is possible characterizing many of the extrinsic and intrinsic muscles during grasp and finding correlations or synergies between muscles will allow for the prediction of intrinsic muscle activity based only the extrinsic signals. This characterization has been done in the past, but without a focus on prediction and more on which muscles contribute to different grasp types (Maier and Hepp-Reymond, 1995).

An ideal solution would be to decode behaviors of the missing interossei directly their original neurons, but multiple studies show that motor neurons degenerate without their original muscle body or without being attached to a new muscle (Carlson et al., 1979; Hoffer et al., 1979; Dhillon et al., 2004). When attached to a new muscle, the signals could be decoded if branches of the nerve are spread like in the case of targeted muscle reinnervation (Kuiken et al., 2017), but this would have to be a decision made at the time of injury.

### Does Peripheral Nerve Stimulation Modify the Error Signal to the Brain?

This study continues our previous work into how stimulation evoked tactile feedback is processed pre-perceptually (Chowdhury and Tyler, 2024) . Previously we showed that PNS is used in the most basic processing pathways at a pre-perceptual level. This study investigated the feedback as a simple error signal to sensorimotor system. Due to limitations to of our prosthesis, the participant could not reduce force, but stimulation seemed to have an instantaneous effect on the movement height of the puck. We hypothesize this instantaneous improvement towards intact like movement is from an update to the internal model, a function of the cerebellum in the brain stem. The stimulation would be used as a pre-perceptual error signal that is compared with the body’s expectation of sensation. However, similar instantaneous improvements have occurred with stimulation evoked tactile feedback without interaction with the brain stem.

Cortical stimulation of the somatosensory cortex also produces instantaneous improvements in motor performance (Flesher et al., 2021) and it is unlikely the cortical stimulation is having a pre-perceptual effect. Previous studies show that cortical stimulation is poorly processed (O’Doherty, 2009; Godlove et al., 2014; Caldwell et al., 2019) and yet the functional improvements are clear for a population of spinal cord injury participants. It is possible that PNS is perceived in the same way as cortical stimulation and providing the same improvements. PNS could provide feedback that the participant uses in understanding how well they are doing, but this is unlikely. In the other studies that showed instantaneous improvements to functional ability, the participants were aware they were performing better given stimulation. In this study, the participant was not aware they were moving the puck any lower in one case verses another. For this reason, these changes still appear to be a pre-perceptual improvement as the participant did not actively choose to move the puck lower depending on what conditions were used in the test. There were consistent and significant differences in movement height which would be difficult to control consciously.

### Comparing Participants with Limb Differences to SCI Participants

Participants with limb differences and survivors of spinal cord injury have vastly different base functional performances due to their differing levels of neural impairment. This makes comparisons of functional improvement difficult. There is arguable a larger gap in functional ability of those with spinal cord injuries (SCI) to able-bodied individuals compared to those with transradial limb differences and able-bodied individuals. The same significant results seen when supplying touch through cortical implants to SCI participants may not apply to those with limb differences. For example, while Flesher saw immediate improvements to functional ability classified as able-bodied performance (movement of an object to a target under 5 seconds) in the ARAT test when cortical stimulation was given vs when it was not (Flesher et al., 2021), the same results would not be seen with our participant. Our participant performed a very similar test to the ARAT in our study and no matter the condition, the participant always completed the movement under 3 seconds. Our participants still have the use of the majority of their limbs which makes this comparison hard and the differences in performance more nuanced.

## Supporting information

Prosthesis Force Controller Validation Data and Sensorized Puck Calibration

## Data Availability

All data produced in the present study are available upon reasonable request to the authors

## Acknowledgments

We thank the DARPA HAPTIX program (N66001-15-4014), the National Institutes of Health (T32AR007505), and the Louis Stokes VA (I01 RX00133401, C3819) for their support of this research. Thank you to Patrick Pariseau and Susan Schramfield for their many hours analyzing and manually labeling video data in DeepLabCut. Without them, I would have no motion capture data. The clinical staff for our research study consisting of Melissa Schmitt, Alexandra Hutchison, Christine Cowen, and Jessica Walrath once again spent countless hours communicating with our participants explaining our studies to them, recruiting for the program, and providing much needed insight into working with a patient population with limb differences. Finally, we would like to thank our participant for dedicating this part of their lives to our study in the hope that their efforts will benefit their greater community.

