## Supplementary material for "Direct Prosthesis Force Control with Tactile Feedback May Connect with the Internal Model": Prosthesis Force Controller Validation Data and Sensorized Puck Calibration

Tuning and Validation of the 1 Dof Force Controller

During the tuning of the 1 DoF force controller in the second major aim of this dissertation, I used the Ziegler–Nichols method (Ziegler and Nichols, 1993) to tune a PID controller for a specific TASKA prosthetic hand (TASKA Prosthetics, Christchurch, New Zealand) to match selected force amplitudes by controlling the hand’s aperture size. The parameters found using the Ziegler-Nicols method are the ultimate gain, $K_{u}$, when a proportional controller gives consistent oscillations and the oscillation period, $T_{u}$. The tuned parameters are $K_{u}=0.1375$ and $T_{u}=7.9167$ and fit into the various controller types below in **Supplementary Table 1**.

*Supplementary Table 1: The various controller types created using parameters from the Ziegler-Nicols method.*

| **Controller Style** | $\boldsymbol{K}_{\boldsymbol{p}}$ | $\boldsymbol{T}_{\boldsymbol{i}}$ | $\boldsymbol{T}_{\boldsymbol{d}}$ | $\boldsymbol{K}_{\boldsymbol{i}}$ | $\boldsymbol{K}_{\boldsymbol{d}}$ |
| --- | --- | --- | --- | --- | --- |
| **P** | $0.5K_{u}$ | $-$ | $-$ | $-$ | $-$ |
| **PI** | $0.45K_{u}$ | $0.8T_{u}$ | $-$ | $0.54\frac{K_{u}}{T_{u}}$ | $-$ |
| **PD** | $0.8K_{u}$ | $-$ | $0.125 T_{u}$ | $-$ | $0.1K_{u}T_{u}$ |
| **PID** | $0.6K_{u}$ | $0.5T_{u}$ | $0.125 T_{u}$ | $1.2\frac{K_{u}}{T_{u}}$ | $0.075K_{u}T_{u}$ |
| **Pessen Integral Rule** | $0.7K_{u}$ | $0.4T_{u}$ | $0.15 T_{u}$ | $1.75\frac{K_{u}}{T_{u}}$ | $0.105K_{u}T_{u}$ |
| **Some overshoot** | $0.3\overline{3}K_{u}$ | $0.5T_{u}$ | $0.3\overline{3}T_{u}$ | $0.66\frac{K_{u}}{T_{u}}$ | $0.1\overline{1}K_{u}T_{u}$ |
| **No overshoot** | $0.2K_{u}$ | $0.5T_{u}$ | $0.3\overline{3}T_{u}$ | $0.4\frac{K_{u}}{T_{u}}$ | $0.6\overline{6}K_{u}T_{u}$ |

Once the PID controller was tuned, it was validated with various input force profiles to analyze its accuracy. These profiles are in **Supplementary Figure 1**, **Supplementary Figure 2**, and **Supplementary Figure 3**.

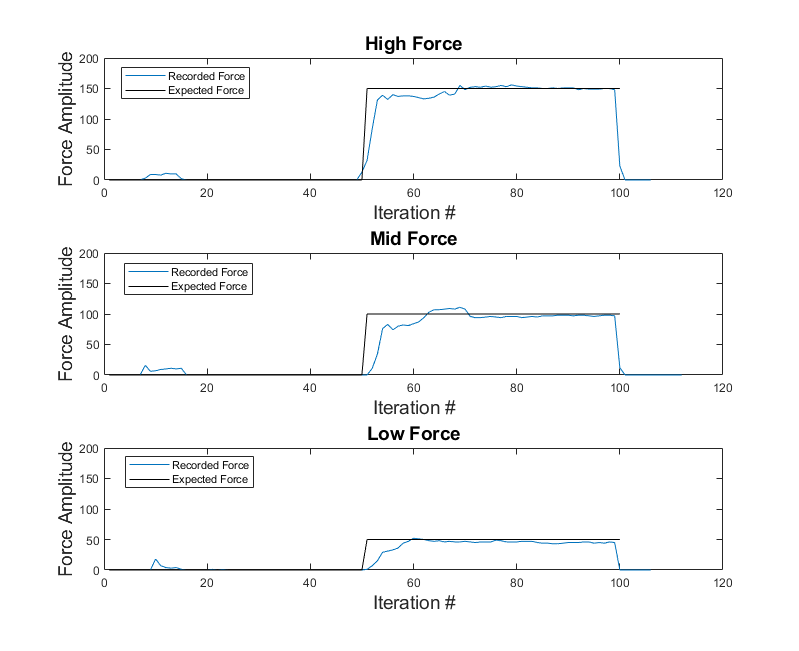

*Supplementary Figure 1: Analysis of the force controller’s response time to a step force input.*

*The Zigler-Nicols method minimizes overshoot in this response. Force was measured by sensors in the TASKA hand’s fingertips.*

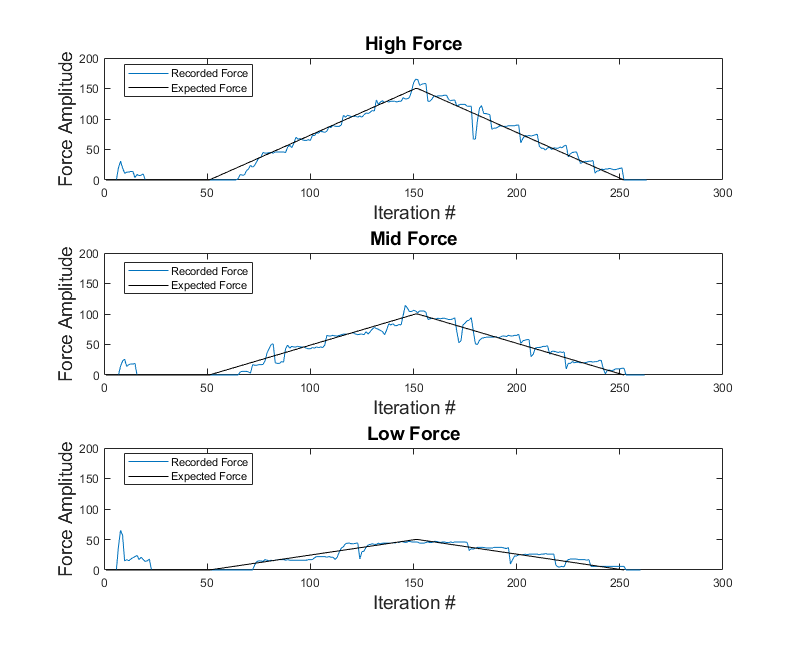

*Supplementary Figure 2: An analysis of the force controller’s response to a ramp input.*

*The TASKA hand could follow the ramp in force well, but due to the low resolution in grip positions, would sometimes oscillate between them. Force was measured by sensors in the TASKA hand’s fingertips.*

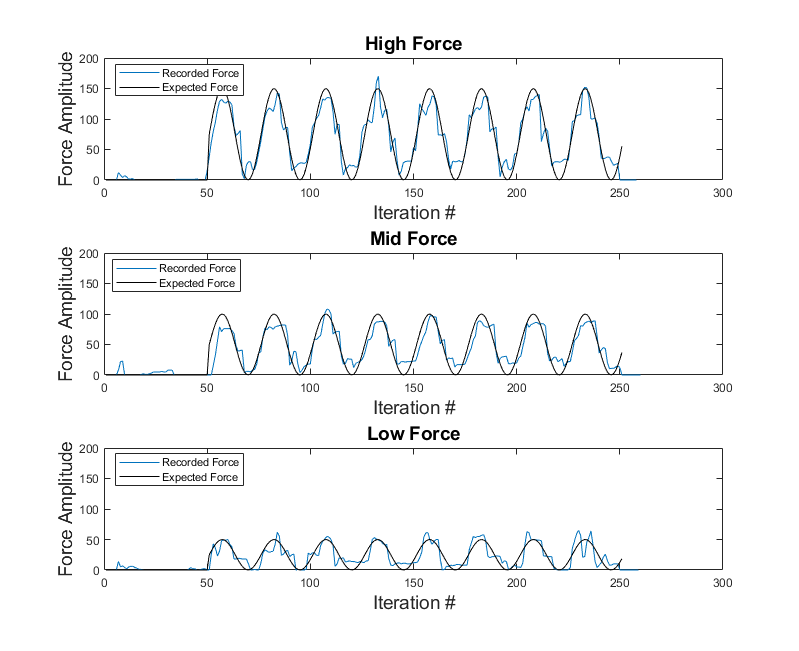

*Supplementary Figure 3: An analysis of the force controller’s response to a sinusoidal input.*

*Much like the ramp response, the force controller followed the commanded force well, but still needed to oscillate between grip positions. Force was measured by sensors in the TASKA hand’s fingertips.*

Conversion of Puck Sensor Data to Force

The FitMi pucks (Flint Rehab, Irvine, CA) report raw values between 0 and 1000 with a baseline near 400 when there is no force on the puck. The conversion of the sensor values to mass on the face of the puck is below in .

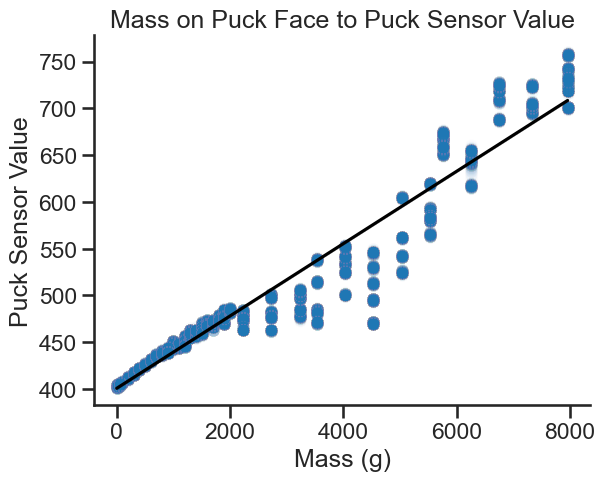

*Supplementary Figure 4: A calibration curve for the sensorized puck used in Chapter 4.*

*The reported sensor values for known masses placed on the face of the puck. Past, 2000 grams, the masses was the combination of the calibrated masses with other heavy objects of measured weights. The large size of these objects led to more wobble on the face of the puck and less consistent measurements.*

The linear fit to the measurements gave an equation of $y=400.8+0.0387x$ with $x$ in grams on the face of the puck. The range of weights tested encompasses the weights participants applied to the puck, but the max sensor value could be achieved given 15,506.2g of mass on the face of the puck. Converting to force on the puck from mass, the puck can measure squeezing forces from 0N to 152.03N.
